# Heterogeneity in Youth Social Media Engagement and Its Pathways to Mental Health and Wellbeing

**DOI:** 10.64898/2026.03.30.26349717

**Authors:** Rui Adele H Wang, Vincent S. Huang, Shanze Sadiq, Peter Smittenaar, Hannah Kemp, Sema K Sgaier

## Abstract

**Introduction:** Social media is a central part of young people’s lives, yet research on its mental health effects remains mixed. We posit that these inconsistencies stem in part from treating youth as a homogeneous group, obscuring distinct behavioral patterns associated with divergent mental health and wellbeing trajectories.

**Objectives:** This study aimed to: (1) explore heterogeneity in social media engagement styles among U.S. youth aged 15-24; and (2) examine how these engagement styles are embedded within a broader system of mental health, wellbeing, emotional regulation, belonging, family and neighborhood context, and stress and adversity.

**Methods:** Data were drawn from a 2024 nationally representative cross-sectional survey of 2,563 U.S. youth, conducted as part of the Youth Mental Health Tracker initiative. We employed unsupervised clustering to identify five distinct social media engagement profiles. Subsequently, we used Bayesian network-based causal discovery to examine (a) upstream factors that emerge as drivers of engagement styles and (b) downstream outcomes influenced by profile membership in the learned system.

**Results:** Five profiles were identified: the Perpetually Plugged-In (31.3%), characterized by near-constant multifaceted social media use, for both positive and negative purposes across multiple domains of life; the Burned-Out Browsers (21.9%), with high exposure to negative and comparison-based content with frequent attempts to disengage; the Practical Navigators (20.7%) who engage in structured, goal-oriented use focused on learning, hobbies, and maintaining connections; the Positive Engagers (13.6%) with high social and identity-driven engagement; and the Light Touch Users (12.5%) who have low overall engagement and limited reliance on social media for connection, identity, or support.

Causal analyses revealed that the Perpetually Plugged-In and Burned-Out Browsers had the worst mental health and wellbeing, with their engagement driven by different reasons. While both engagement profiles were influenced by similar psychosocial risk factors, they were distinguished by their dominant drivers: contemporaneous social stressors (bullying, discrimination, and emotional dysregulation) for Perpetually Plugged-In youth, versus adverse childhood experiences for Burned-Out Browsers. In contrast, Positive Engagers reported high social media engagement alongside the highest levels of social wellbeing, using social media for identity exploration and social support within a context of low cumulative stress and adversity.

**Conclusions:** Findings suggest that youth social media risk is not driven by intensity of use alone, but by the interaction between engagement style and offline emotional and social conditions. Policies focused solely on restricting access risk overlooking these differences and may inadvertently sever important sources of connection for many youth. Strategies should identify experiential risk signals while strengthening supportive contexts that enable healthier engagement. Overall, youth social media use is best understood as part of a broader psychosocial system, and recognizing this heterogeneity is essential for designing more targeted, equitable, and evidence-based interventions.

## Introduction

Social media has become a central part of young people’s daily lives, shaping how they connect, learn, and express themselves. A considerable amount of the existing literature focuses on the problematic or “addictive” aspects of social media use (SMU) (Bekalu et al., 2022; Finserås et al., 2023; Peng & Liao, 2023; Schivinski et al., 2020), leaving less attention to the ways young people may engage with social media in adaptive, positive, or prosocial ways (Vaingankar et al., 2022). Recently, there are increasingly nuanced conceptualizations of social media engagement, moving beyond one-dimensional measures of “screen-time” toward examining what young people are using social media for (Valkenburg et al., 2022). Studies find mixed associations between SMU and mental health and wellbeing outcomes (Bekalu et al., 2022; Orben, 2020; Teague et al., 2026; Valkenburg et al., 2022), with some studies emphasizing risks linked to problematic or excessive use (Shannon et al., 2022; Shieun Lee et al., 2022), others finding null effects (Cheng et al., 2026) and others still identifying benefits related to social connection, identity exploration, learning, and access to support (Marciano & Viswanath, 2023; van der Wal et al., 2024).

A growing body of research underscores the nuanced nature of SMU, showing that its effects on youth mental health and wellbeing depend heavily on how, and in what contexts, young people use these platforms (Agyapong-Opoku et al., 2025; Fassi et al., 2025; Valkenburg et al., 2022). Social media experiences unfold within social, cultural, and contextual environments, and social media practices fit within offline lives that shape how online engagement translates into mental health and wellbeing outcomes. At the individual level, engagement style, emotion regulation, and resilience may influence whether online engagement is experienced as supportive or stressful (Boer et al., 2021; Piccerillo & Digennaro, 2025). At the social level, family acceptance and peer belonging can buffer the negative impacts of social media experiences (Abebe et al., 2024; Meshi & Ellithorpe, 2021). At the structural level, exposure to discrimination and marginalization may further condition the effects of SMU (Jackson et al., 2021; Tao & Fisher, 2023). Viewed through this multidimensional lens, social media may function less as a monolithic determinant of mental health and more as an amplifier of existing psychological, social, and material resources.

Although research on youth and digital media has expanded rapidly in recent years, U.S. population-level analyses that examine multidimensional patterns of digital media use within youths’ broader social and developmental contexts remain sparse. Using 2018 National Survey of Children’s Health data, Jackson et al. (2021) found that heavy digital media use was strongly associated with cumulative adverse childhood experiences, with family resilience, family connection, and parenting stress mediating the association. While foundational, the study relied on coarse measures of digital engagement and a limited set of contextual factors. Given the rapid evolution of social media platforms, youth online behaviors and policies and legislations, there is a critical need for updated, multidimensional analyses that capture how contemporary patterns of social media use intersect with social context, wellbeing, and mental health among U.S. youth. One-size-fits-all policies or interventions may be suboptimal when risks and benefits are unevenly distributed across youth. Segmentation analysis offers a valuable, data-driven first step toward addressing this complexity by identifying naturally occurring subgroups defined by shared patterns of social media use, offline context, and psychosocial functioning, providing a foundation for more targeted research, intervention, and policy design (Keum et al., 2023; Winstone et al., 2022).

While segmentation captures heterogeneity in youth social media engagement, it remains descriptive and cannot, on its own, distinguish correlates from intervention-relevant mechanisms (Orben, 2020). Moving from descriptive typologies toward hypothesis generation about plausible mechanisms is critical for informing intervention, as improving youth outcomes requires not only identifying at-risk groups but understanding which factors, if changed, may alter those outcomes. To this end, we apply causal discovery methods that learn directed acyclic graphs (DAGs) from observational data, representing conditional dependencies among variables as causal Bayesian networks (BNs; Holmes & Jain, 2008). We use these models to (i) propose a plausible system structure consistent with the observed data and pre-specified temporal constraints, and (ii) conduct model-based “what-if” simulations that infer how changes in upstream factors would be expected to shift the probability of different engagement profiles and downstream outcomes.

This study integrates multidomain measures of social media engagement, mental health, wellbeing, emotional regulation, belonging, family and neighborhood context, and exposure to stress and adversity using a nationally representative sample of US youth. We identify distinct social media engagement personas and, by combining segmentation with model-based causal analysis, examine (a) upstream factors that emerge as drivers of engagement styles and (b) downstream outcomes influenced by segment membership in the learned system. Together, these analyses offer a systems-level population-level portrait and a set of prioritized, testable hypotheses to inform more targeted, context-sensitive public health and digital wellbeing strategies for diverse youth populations.

## Results

### Sample Characteristics

Table 1 and 2 shows key demographics of the sample, variables used for the causal analysis across the sample, and the distribution of the segmentation variables.

**Table 1.** Variables for causal analysis, with prevalence in youth ages 15-24.

|  | Percentage | Confidence Intervals |
| --- | --- | --- |
| Satisfied with life overall (Very satisfied or satisfied) | 81.60% | 79.2%, 83.7% |
| Reporting symptoms of depression (PHQ2) and/or anxiety (GAD2) | 30.50% | 27.9%, 33.3% |
| Struggled with mental health in the past 2 years | 44.40% | 41.5%, 47.3% |
| Overall, I feel that what I do in my life is meaningful (Slightly or strongly agree) | 82.20% | 79.9%, 84.3% |
| Usually feel a sense of accomplishment from what I do (Slightly or strongly agree) | 84.40% | 82.1%, 86.4% |
| There is a person with whom I can share joys and sorrows (Slightly or strongly agree) | 88.20% | 86.2%, 90.0% |
| I get the emotional help and support I need from my friends (Slightly or strongly agree) | 80.60% | 78.1%, 82.8% |
| I get the emotional help and support I need from my family (Slightly or strongly agree) | 78.00% | 75.5%, 80.3% |
| I feel like I belong as part of a group, like my friends, school, or where I live (Slightly or strongly agree) | 76.80% | 74.3%, 79.2% |
| My family accepts me (Slightly or strongly agree) | 89.00% | 86.9%, 90.7% |
| I get along well with people my age (Slightly or strongly agree) | 86.50% | 84.4%, 88.4% |
| I feel like I can do things to help others, such as in my family or with my friends, volunteering in my community, or giving to charity (Slightly or strongly agree) | 87.10% | 85.0%, 88.9% |
| In the last month, I felt difficulties were piling up so high that I could not overcome them (Slightly or strongly agree) | 44.90% | 42.0%, 47.9% |
| I find it hard to let go of bad feelings (Slightly or strongly agree) | 62.10% | 59.2%, 64.9% |
| I find it hard to express myself to others (Slightly or strongly agree) | 60.70% | 57.7%, 63.6% |
| I am confident that I can achieve what I set out to do (Slightly or strongly agree) | 84.30% | 82.0%, 86.4% |
| When things go wrong in my life I usually get back to normal quickly (Slightly or strongly agree) | 73.60% | 70.9%, 76.1% |
| In the past year I have been bullied or cyberbullied (Slightly or strongly agree) | 22.40% | 20.1%, 24.9% |
| Experienced any adverse childhood experiences | 51.00% | 48.1%, 54.0% |
| If I had a mental health problem, I would be embarrassed to tell other people (Slightly or strongly agree) | 48.00% | 45.0%, 51.0% |
| Experienced any identity based discrimination | 48.60% | 45.7%, 51.6% |
| Physical health (Good or very good) | 90.70% | 88.9%, 92.3% |
| I feel safe in my neighborhood (Slightly or strongly agree) | 87.30% | 85.4%, 89.0% |
| People in my neighborhood look out for each other (Slightly or strongly agree) | 66.80% | 63.9%, 69.5% |
| Worried about any global issues | 83.50% | 81.1%, 85.7% |
| Worried about any social issues | 79.50% | 76.9%, 82.0% |
| Age: 15-17 | 41.40% | 38.5%, 44.4% |
| Age: 18-24 | 58.60% | 55.6%, 61.5% |
| Gender: Another | 2.20% | 1.55%, 3.16% |
| Gender: Man | 49.20% | 46.3%, 52.2% |
| Gender: Woman | 48.60% | 45.6%, 51.5% |
| Race/Ethnicity: Asian | 5.80% | 4.54%, 7.35% |
| Race/Ethnicity: Black | 13.40% | 11.7%, 15.4% |
| Race/Ethnicity: Hispanic | 24.30% | 21.9%, 26.9% |
| Race/Ethnicity: Other | 5.40% | 4.28%, 6.80% |
| Race/Ethnicity: White | 51.00% | 48.1%, 54.0% |
| LGBTQ (vs non-LGBTQ+) | 21.40% | 19.0%, 23.9% |
| Receive any government assistance (vs did not receive) | 29.70% | 27.3%, 32.3% |
| Living in metropolitan area (vs non-metropolitan area) | 87.80% | 85.8%, 89.6% |
| Living with caregivers | 75.30% | 72.7%, 77.6% |

**Table 2.** Social media engagement variables used for segmentation, with prevalence in youth ages 15-24.

|  | Percentage | Confidence Intervals |
| --- | --- | --- |
| Social media makes me feel bad about my appearance sometimes or all the time | 46.10% | 43.3%, 48.9% |
| Social media makes me feel not as good as other sometimes or all the time | 50.00% | 47.2%, 52.8% |
| Social media makes me feel bad about the world sometimes or all the time | 70.10% | 67.4%, 72.7% |
| I use digital devices like mobile phones, laptops and gaming devices almost constantly every day (slightly or strongly agree) | 84.00% | 81.8%, 86.0% |
| I have tried to reduce the amount of time I spend on digital devices (slightly or strongly agree) | 62.30% | 59.5%, 65.0% |
| Social media helps me learn about issues in the world sometimes or all the time | 78.10% | 75.8%, 80.3% |
| Social media is important for me to explore my identity (slightly or strongly agree) | 48.00% | 45.1%, 50.8% |
| Social media is important for me to connect with my friends (slightly or strongly agree) | 76.90% | 74.4%, 79.2% |
| Social media is important for me in my hobbies or interests (slightly or strongly agree) | 69.30% | 66.6%, 71.8% |
| Social media is important for me to give and get emotional support to and from others (slightly or strongly agree) | 50.70% | 47.9%, 53.5% |
| Have tried spending less time on social media for my mental health | 83.50% | 81.2%, 85.6% |

### Segmentations

Weighted k-medoids clustering was evaluated across k = 2-8 using total weighted within-cluster distance and average silhouette width. While within-cluster distance declined monotonically with increasing k, average silhouette width peaked at k = 5 (0.27), indicating the best balance between within-cluster cohesion and between-cluster separation (Supplementary 1). Modest absolute silhouette values are consistent with behavioral data (Ranjan & Kumar, 2025). Accordingly, a five-segment solution was selected. Table 3 details the 5 social media engagement segments that emerged. Segments were named to reflect their key differentiating social media use characteristics.

**Table 3.** Social media engagement segments descriptions and mental health and wellbeing outcomes.

| Segmentation variables | Perpetually Plugged-In (n = 779; 31.3%) |  | Burned-Out Browsers (n = 547; 21.9%) |  | Practical navigators (n = 516; 20.7%) |  | Positive engagers (n = 339; 13.6%) |  | Light touch users (n = 312; 12.5%) |  |
| --- | --- | --- | --- | --- | --- | --- | --- | --- | --- | --- |
| Description | Near-constant device use with social media central to connection, identity, and support. |  | High exposure to negative and comparison-based content with frequent attempts to disengage. |  | Structured, goal-oriented use focused on learning, hobbies, and maintaining connections. |  | High social and identity-driven engagement, paired with active efforts to reduce use. |  | Low overall engagement, limited reliance on social media for connection, identity, or support. |  |
| I see things on social media that make me feel bad about my appearance or body | 84.50% | 80.5%,<br>87.8% | 74.60% | 68.8%,<br>79.7% | 5.20% | 3.27%,<br>8.25% | 8.70% | 5.44%,<br>13.5% | 9.70% | 5.94%,<br>15.5% |
| I see things on social media that make me feel like I am not as good as other people | 87.50% | 83.7%,<br>90.4% | 81.20% | 76.0%,<br>85.5% | 13.50% | 9.45%,<br>18.8% | 6.90% | 4.18%,<br>11.2% | 10.70% | 6.59%,<br>16.8% |
| I see things on social media that make me feel unhappy about the world | 94.90% | 92.5%,<br>96.5% | 91.90% | 87.8%,<br>94.7% | 71.50% | 65.0%,<br>77.2% | 20.00% | 14.6%,<br>26.8% | 22.10% | 16.1%,<br>29.5% |
| I use digital devices like mobile phones, laptops and gaming devices almost constantly every day | 92.10% | 88.7%,<br>94.5% | 82.60% | 77.8%,<br>86.4% | 84.00% | 78.6%,<br>88.2% | 87.50% | 81.1%,<br>91.9% | 63.80% | 55.2%,<br>71.5% |
| I have tried to reduce the amount of time I spend on digital devices | 67.40% | 62.5%,<br>71.9% | 74.30% | 68.7%,<br>79.2% | 31.40% | 25.9%,<br>37.5% | 79.00% | 72.2%,<br>84.6% | 59.30% | 50.8%,<br>67.4% |
| Tried spending less time on social media | 86.60% | 82.7%,<br>89.7% | 90.50% | 86.0%,<br>93.7% | 71.70% | 65.3%,<br>77.2% | 86.60% | 80.0%,<br>91.2% | 80.10% | 72.9%,<br>85.7% |
| Spending less time on social media was helpful | 61.70% | 56.5%,<br>66.6% | 63.80% | 57.5%,<br>69.6% | 44.70% | 37.4%,<br>52.2% | 61.20% | 52.9%,<br>68.8% | 65.10% | 55.7%,<br>73.5% |
| Social media helps me learn about issues in the world | 91.40% | 88.4%,<br>93.7% | 84.50% | 80.0%,<br>88.1% | 81.80% | 76.4%,<br>86.1% | 78.80% | 72.3%,<br>84.1% | 27.10% | 20.3%,<br>35.1% |
| Social media is important for me in my hobbies or interests | 93.20% | 90.2%,<br>95.4% | 34.80% | 29.4%,<br>40.7% | 91.60% | 87.4%,<br>94.5% | 90.90% | 85.9%,<br>94.3% | 11.50% | 7.41%,<br>17.4% |
| Social media is important for me to explore my identity | 87.20% | 84.0%,<br>89.8% | 6.40% | 3.95%,<br>10.1% | 28.60% | 23.1%,<br>34.8% | 91.50% | 87.2%,<br>94.5% | 8.10% | 5.04%,<br>12.8% |
| Social media is important for me to connect with my friends | 88.90% | 85.2%,<br>91.7% | 69.40% | 63.6%,<br>74.7% | 84.00% | 77.8%,<br>88.7% | 95.60% | 92.5%,<br>97.4% | 28.10% | 21.2%,<br>36.4% |
| Social media is important for me to give and get emotional support to and from others | 83.90% | 80.1%,<br>87.0% | 12.00% | 8.71%,<br>16.3% | 31.30% | 25.9%,<br>37.2% | 96.40% | 93.0%,<br>98.2% | 18.60% | 12.7%,<br>26.4% |
| Flourish: Overall, I feel that what I do in my life is meaningful | 79.90% | 75.8%,<br>83.4% | 75.70% | 70.3%,<br>80.3% | 82.40% | 77.1%,<br>86.7% | 92.70% | 87.9%,<br>95.6% | 86.40% | 80.2%,<br>90.9% |
| Satisfied with life | 74.60% | 70.2%,<br>78.6% | 77.40% | 72.3%,<br>81.9% | 84.00% | 78.6%,<br>88.2% | 94.10% | 90.2%,<br>96.5% | 88.10% | 81.9%,<br>92.3% |
| Symptoms of depression and/or anxiety | 39.50% | 34.9%,<br>44.3% | 37.90% | 32.3%,<br>43.9% | 21.70% | 16.8%,<br>27.4% | 16.70% | 11.9%,<br>22.9% | 23.40% | 16.9%,<br>31.4% |
| Significant mental health struggles in past 2 years | 58.10% | 53.1%,<br>62.9% | 58.50% | 52.2%,<br>64.4% | 31.50% | 25.8%,<br>37.9% | 22.50% | 16.9%,<br>29.3% | 27.20% | 20.2%,<br>35.5% |

Two high risk segments emerge: the Perpetually Plugged-In (n = 779; 30%) and Burned-Out Browsers (n = 547; 21%). Both segments report the worst mental health and lowest wellbeing. Although the two segments both experience high exposure to negative comparisons, their engagement patterns differ sharply in intensity and function. Perpetually Plugged-In youth engage intensely across multiple domains of life, for both positive and negative purposes, whereas Burned-Out Browsers exhibit more constrained, less functional use alongside efforts to disengage. Perpetually Plugged-In youth reported more constant device use (92.1%, 95% CI: 88.7-94.5 vs. 82.6%, 77.8%-86.4%) and were more likely to view social media as central to multiple domains of daily life. Nearly all reported using social media for hobbies or interests (93.2%, 90.2%-95.4%), compared with just over one-third of Burned-Out Browsers (34.8%, 29.4%-40.7%). Differences were even more pronounced for identity- and relationship-based uses: 87.2% (84.0%-89.8%) of Perpetually Plugged-In youth said social media was important for exploring their identity, versus only 6.4% (4.0%-10.1%) of Burned-Out Browsers, and 83.9% (80.1–87.0) relied on it for emotional support, compared with 12.0% (8.7%-16.3%).

Practical Navigators (n = 516; 20%) demonstrated more instrumental and goal-oriented use. While most used social media for learning (81.8%, 76.4%-86.1%) or hobbies (91.6%, 87.4%-94.5%), only a small minority reported comparison-driven harms, such as feeling worse about their appearance (5.2%, 3.3%-8.3%). Positive Engagers (n = 339; 13%) combined high social and identity-driven use: over 90% used social media to connect with friends (95.6%, 92.5%-97.4%) and explore identity (91.5%, 87.2%-94.5%), with strong self-regulation, as nearly four in five reported efforts to reduce device use (79.0%, 72.2%-84.6%). Despite heavy engagement, their mental health and wellbeing profiles were among the strongest, with low levels of depression or anxiety (16.7%, 11.9%-22.9%) and high life satisfaction (94.1%, 90.2%-96.5%). Light-Touch Users (n = 312; 12%) reported the lowest overall engagement, minimal exposure to negative social comparison (9.7% feeling worse about appearance; 5.9%-15.5%), and comparatively low prevalence of symptoms of depression and/or anxiety (23.4%, 16.9%-31.4%).

### Causal structure and upstream drivers of segment membership

A Bayesian network incorporating 34 variables was learned to characterize conditional dependencies linking emotional regulation, adversity, social context, demographics, and segment membership (Supplementary 2). Three factors emerged as direct upstream drivers of segment assignment: difficulty managing negative emotions, experiences of bullying, and exposure to adverse childhood experiences (ACEs). Demographic characteristics (age, LGBTQ identity, and race/ethnicity) and broader social context were positioned upstream in the learned network, influencing segment membership indirectly through pathways involving ACE exposure, discrimination, bullying, and emotional regulation.

**Figure 1.**
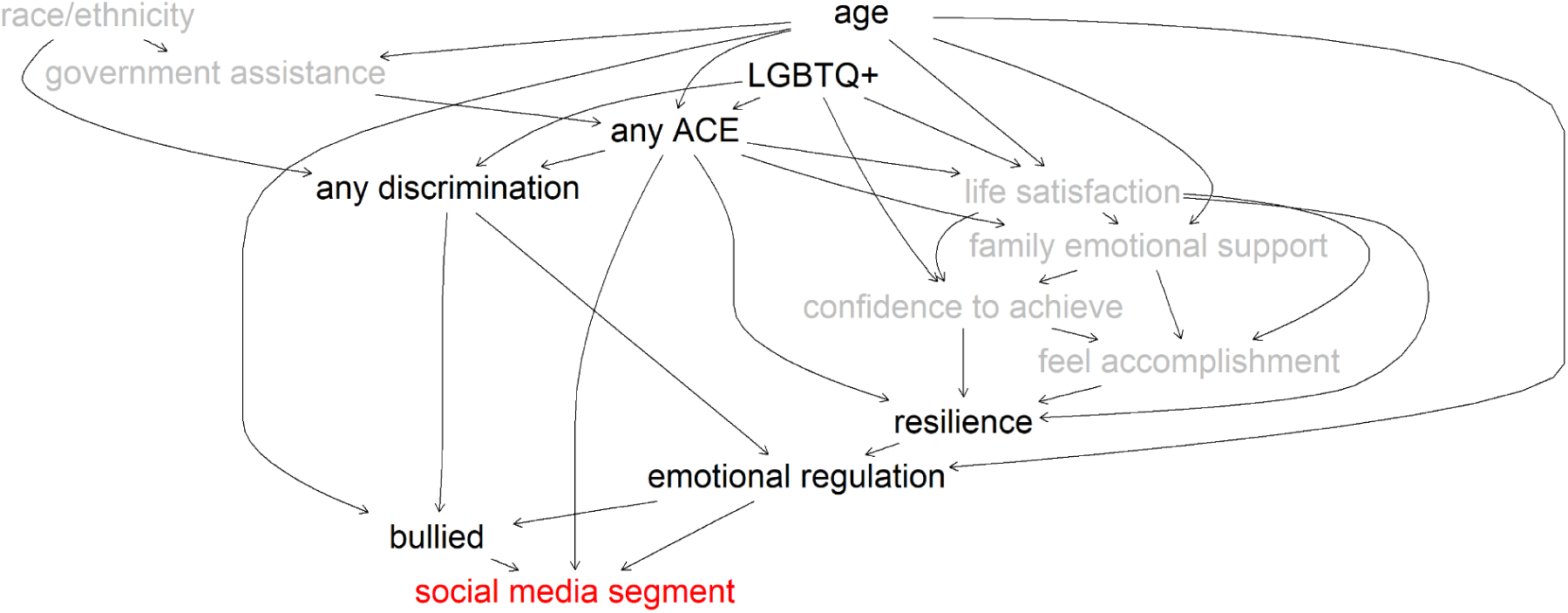
The causal Bayesian Network structure, depicted as a directed acyclic graph (DAG). For clarity, we only include nodes that are direct (immediately upstream) or indirect causes (further upstream) of social media segment membership.

#### Model-based pathways into higher-risk engagement styles

Causal inference analysis revealed asymmetries in how engagement segments change under counterfactual interventions on upstream factors (Supplementary 3). Intervention analyses revealed that transitions into higher-risk social media engagement styles were consistently driven by increases in emotional strain, cumulative adversity, and social threat, regardless of the lower- or moderate-risk segment used as the reference. Across specifications, worsening emotional regulation, exposure to adverse childhood experiences (ACEs), bullying, and discrimination systematically increased the odds of shifting into both Burned-Out Browsing and Perpetually Plugged-In engagement patterns.

When Positive Engagers were used as the reference segment, increased difficulty letting go of negative feelings was associated with higher simulated odds of shifting into Burned-Out Browsers (OR = 2.30, 95% CI: 1.88–2.81) and Perpetually Plugged-In users (OR = 3.29, 95% CI: 2.72–3.98). Similarly, ACE exposure was associated with higher odds of shifting into Burned-Out Browsing (OR = 3.63, 95% CI: 2.94–4.47) and Perpetually Plugged-In use (OR = 2.79, 95% CI: 2.31–3.38). Reductions in the difficulty bouncing back when things go wrong also increased the likelihood of entering both high-risk segments (Burned-Out Browsers: OR = 1.30, 95% CI: 1.07–1.59; Perpetually Plugged-In: OR = 1.34, 95% CI: 1.11–1.61). Bullying exposure increased the odds of shifting into Perpetually Plugged-In use (OR = 1.35, 95% CI: 1.12–1.61). In contrast, bullying was associated with lower odds of shifting into Burned-Out Browsing (OR = 0.67, 95% CI: 0.55–0.82).

Across all reference segments, Burned-Out Browser was most driven by childhood adverse experiences, whereas Perpetually Plugged-In use driven by to contemporaneous social stressors. When Burned-Out Browsers were used as the reference group, increases in current social threat substantially raised the odds of transitioning into Perpetually Plugged-In use, including exposure to bullying (OR = 1.97, 95% CI: 1.67–2.31), difficulty managing negative emotions (OR = 1.42, 95% CI: 1.20–1.68), and experiences of discrimination (OR = 1.19, 95% CI: 1.02–1.40). In contrast, when Perpetually Plugged-In youth served as the reference, the presence of adverse childhood experiences increased the odds of transitioning into Burned-Out Browsing (OR = 1.19, 95% CI: 1.01–1.40).

#### Demographic differences

Age and LGBTQ+ identity were drivers with higher simulated odds of shifting from lower- and moderate-risk engagement styles into higher-risk segments across model specifications. These associations were positioned upstream in the learned network and were largely mediated through bullying exposure, discrimination, and emotional regulation variables. When Positive Engagers were used as the reference group, older youth (ages 18–24) had significantly higher odds of transitioning into Burned-Out Browsers (OR = 1.49, 95% CI: 1.22–1.81) and Perpetually Plugged-In users (OR = 1.36, 95% CI: 1.13–1.63), with similar patterns observed for LGBTQ+ youth (Burned-Out Browsers: OR = 1.36, 95% CI: 1.11–1.66; Perpetually Plugged-In: OR = 1.37, 95% CI: 1.13–1.65). Parallel effects were observed when Light-Touch Users served as the reference, where both older age and LGBTQ+ identity increased the likelihood of transitioning into Burned-Out Browsing (age OR = 1.41, 95% CI: 1.14–1.74; LGBTQ+ OR = 1.27, 95% CI: 1.02–1.57) and Perpetually Plugged-In use (age OR = 1.36, 95% CI: 1.11–1.61; LGBTQ+ OR = 1.33, 95% CI: 1.09–1.63).

Race and ethnicity were also positioned upstream of engagement styles within the learned causal structure, but their effects on segment transitions were largely indirect. Similarly, receipt of government assistance lay on the causal pathway to segment membership but was not itself a strongly significant direct driver of transitions. In the learned network, both race/ethnicity and government assistance were strongly linked to adverse childhood experiences and exposure to discrimination, which in turn exerted direct causal influence on engagement style transitions.

### Impact of segment membership on psychobehavioral outcomes

Downstream (Figure 2, Supplementary 4) intervention analyses showed a clear gradient in adverse outcomes, with the strongest downstream shifts concentrated in the Perpetually Plugged-In and Burned-Out Browsers segments. Relative to remaining Positive Engagers, shifting youth into Burned-Out Browsers causes higher simulated odds of adverse downstream outcomes, including substantially higher odds of lower perceived ability to help others (OR = 1.55, 95% CI: 1.25–1.92), lower belonging (OR = 2.56, 2.14–3.07), having struggled with mental health in the past two years OR = 1.94, 1.70–2.23), greater mental health stigma (OR = 2.11, 1.87–2.39) and elevated stress (OR = 1.20, 1.06–1.37), alongside higher odds of depression and anxiety symptoms (OR = 1.46, 1.24–1.73). Assignment to Perpetually Plugged-In also showed an even more pronounced downstream profile, with large increases in the odds of reporting depression/anxiety symptoms (OR = 2.25, 1.92–2.64), having experienced mental health struggles in the past two years (OR = 2.79, 2.44–3.19), mental health stigma (OR = 2.54, 2.25–2.87), high stress (OR = 2.13, 1.88–2.41), low belonging (OR = 1.70, 1.41–2.04), and reduced perceived capacity to help others (OR = 1.50, 1.21–1.86). In contrast, shifts from Positive Engagers into Practical Navigators were mixed, with higher odds of low belonging (OR = 1.45, 1.20–1.76) and reduced perceived ability to help others (OR = 1.45, 1.15–1.81), but lower odds of depression/anxiety symptoms (OR = 0.68, 0.56–0.82) and stress (OR = 0.60, 0.52–0.70). Moves from Positive Engagers into Light-Touch Users were comparatively modest and, for some outcomes, protective, including lower odds of recent mental health struggles (OR = 0.71, 95% CI: 0.61–0.83) and stress (OR = 0.79, 95% CI: 0.69–0.91). However, this shift was also associated with a small but significant increase in the odds of lower perceived belonging (OR = 1.23, 95% CI: 1.01–1.50), suggesting that reduced engagement may alleviate distress for some youth while simultaneously weakening feelings of social connection.

**Figure 2.**
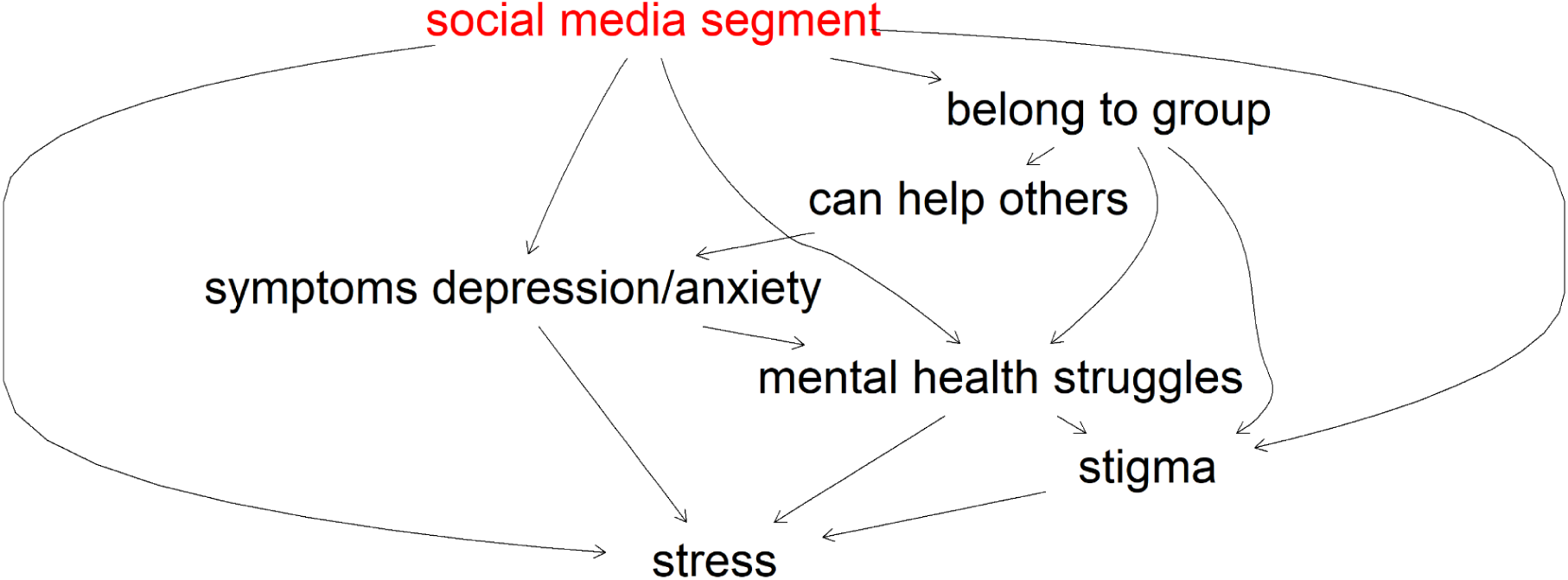
Downstream factors from social media segments.

## Discussion

This study provides a contemporary, population-level portrait of how U.S. youth engage with social media and how these engagement styles are embedded within broader systems of emotional regulation, social context, and cumulative adversity. Rather than examining isolated platform behaviors or screen time measures, we identify five distinct engagement styles that reflect how youth integrate social media into identity development, relationships, learning, and emotional coping. Two segments, Perpetually Plugged-In and Burned-Out Browsers, emerge as particularly vulnerable, accounting for roughly half of youth in the sample and exhibiting substantially worse mental health and wellbeing.

Our findings advance existing research in two important ways. First, the segmentation analysis demonstrates that problematic social media experiences are not evenly distributed across youth, but instead cluster into distinct behavioral profiles that combine intensity of use, motivations for engagement, and exposure to negative comparison or distressing content. Importantly, the highest-risk groups were not defined simply by heavy usage. While Perpetually Plugged-In youth reported near-constant engagement and relied heavily on social media for identity exploration and emotional support, Burned-Out Browsers exhibited lower functional engagement yet similarly high exposure to negative comparison and distress. In contrast, Positive Engagers reported comparably high levels of engagement for connection, identity exploration, and emotional support but showed substantially stronger wellbeing profiles and far lower exposure to comparison-driven harms. This contrast underscores that intensity of use alone does not determine risk.

Second, by integrating segmentation with causal Bayesian network analysis, the study shows that the emotional and social contexts shaping how youth engage with social media play a central role in whether digital environments function as sources of support or amplifiers of distress. Model-based intervention analyses suggested that increases in emotional dysregulation, social threat, and cumulative adversity raised the likelihood of transitions into the two high-risk engagement styles, whereas youth embedded in more supportive contexts, characterized by stronger emotion regulation, and lower exposure to bullying or discrimination, were more likely to exhibit the Positive Engager profile. Together, these findings indicate that differences in youth social media experiences are less about how much young people use digital platforms and more about the emotional and social conditions that shape how those platforms are used (Odgers&Jensen, 2020, Casteele et al., 2024). Importantly, the causal network also positioned segment membership upstream of mental health and wellbeing outcomes, showing that the way young people engage with digital platforms actively influences their broader mental health and wellbeing trajectories.

While prior work has highlighted a wide range of individual, social, and structural factors that may shape how youth experience social media, not all of these influences appeared as direct drivers of engagement style in our causal model. Instead, the learned network revealed a hierarchical structure of influence in which more distal contextual conditions operate through a smaller set of proximal mechanisms. Factors such as family support, peer belonging, and socioeconomic context were present in the network but were typically positioned further upstream or downstream rather than directly determining segment membership. Structural and demographic conditions, including discrimination, race/ethnicity, and socioeconomic disadvantage, primarily operated through pathways involving adversity and social stress. This layered structure suggests that social media engagement styles may emerge from cascading influences across levels of young people’s environments. This hierarchical perspective may also help explain why prior studies have often found inconsistent or weak associations between demographic and contextual factors and social media outcomes (Bekalu et al., 2019, Godard & Holtzman, 2024): many of these influences operate indirectly through intermediate emotional and social processes rather than directly shaping digital behavior.

### Social media engagement as a reflection of offline context

Perpetually Plugged-In youth are characterized by near-constant device use, with social media occupying a central role in social connection, identity exploration, and emotional support. Our model-based analyses prioritize contemporaneous social and emotional strain as plausible antecedents of this profile: difficulty managing negative emotions, exposure to bullying, and experiences of discrimination were consistently positioned proximal to segment membership. We interpret these patterns as consistent with a compensatory hypothesis in which intensive engagement may function as a coping response to immediate social threat and emotional distress (Wolfers&Utz, 2022). When offline environments feel unsafe, invalidating, or insufficiently supportive, digital spaces may offer more accessible avenues for connection, affirmation, and emotional expression. However, reliance on social media for core psychosocial needs may also increase exposure to comparison, conflict, and emotionally charged content, potentially reinforcing cycles of stress and distress (Wolfers&Utz, 2022, Shannon et al., 2022, Marino 2018). In this way, Perpetually Plugged-In engagement appears less as excessive use per se and more as an adaptive, but potentially costly, response to ongoing social and emotional challenges.

Burned-Out Browsers represent a distinct high-risk engagement style characterized by persistent exposure to negative and comparison-driven content alongside markedly lower functional reliance on social media for connection, identity, or emotional support. Adverse childhood experiences emerged as the dominant causal driver of Burned-Out Browsing (Crouch et al., 2025), suggesting that this pattern reflects longer-term emotional exhaustion rather than an acute coping response. Youth in this segment frequently reported attempts to reduce or disengage from social media, yet continued to encounter distressing content, indicating a form of passive, fatigued engagement rather than active seeking of connection or support. This profile is consistent with a depletion model, in which prolonged exposure to stress undermines emotional recovery and motivation, leaving youth more vulnerable to negative affective spillover from digital environments even as they attempt to withdraw (Sommerville 2014, Barke et al., 2024, Reineike et al., 2014). In this context, social media may function less as a compensatory resource and more as a residual stressor that youth feel unable to fully escape, reinforcing cycles of disengagement, cynicism, and diminished wellbeing.

When youth are treated as a homogeneous group and social media exposure is operationalized primarily as time spent online, fundamentally different engagement styles with opposing implications are collapsed into a single estimate, which may explain the small and mixed patterns in social media studies (Orben 2020, Valkenburg et al., 2022). In our data, high-intensity social media use characterizes both youth with the poorest outcomes (Perpetually Plugged-In) and those with some of the strongest wellbeing profiles (Positive Engagers). The Positive Engagers segment demonstrates that high engagement with social media is not inherently detrimental. Youth in this group reported intensive use for connection, identity exploration, and emotional support, often at levels comparable to Perpetually Plugged-In youth, yet exhibited among the strongest mental health and wellbeing profiles.

Causal analyses further suggest that Positive Engagers are not simply “high users who got lucky,” but are embedded in more supportive emotional and social contexts. Relative to high-risk segments, Positive Engagers experienced lower exposure to bullying and discrimination, stronger emotion regulation capacities, and higher levels of belonging and perceived support. These upstream conditions appear to enable youth to use social media as a resource rather than a refuge, extending existing strengths rather than compensating for acute or chronic deficits. Downstream intervention analyses reinforce this distinction: shifts into Positive Engager profiles were associated with substantially lower odds of depression and anxiety symptoms, reduced stress, and higher life satisfaction and belonging compared with transitions into high-risk segments, while remaining comparable, or in some cases superior, to Light-Touch use (Smith et al., 2021, Dredge&Schreurs, 2020). Together, these findings underscore that the benefits or harms of social media are driven less by intensity of use than by the emotional and social contexts that shape how youth engage. Because these downstream patterns are derived from observational BN simulations, they should be interpreted as priorities for prospective testing (e.g., longitudinal tracking of within-person transitions between engagement profiles and quasi-experimental evaluation of upstream levers such as bullying reduction and emotion regulation supports).

#### Implications for intervention and policy

These findings carry important implications for public health, education, and digital wellbeing initiatives. Given the growing wave of legislation restricting or banning youth social media use, these findings underscore the need for more nuanced, evidence-based policy approaches. Rather than treating youth social media use as uniformly harmful, this study highlights how risks and benefits depend on patterns of engagement and offline context, suggesting that future research can help inform policies that are more targeted, proportionate, and responsive to the heterogeneous realities of young people’s digital lives.

Interventions should move beyond blanket recommendations to reduce screen time and instead focus on identifying experiential risk signals, such as comparison-driven distress, bullying exposure, and emotional dysregulation (McAlister et al., 2024) that differentiate harmful from adaptive engagement. Second, prevention strategies should be tailored to distinct causal profiles. Programs that reduce peer victimization and strengthen emotion regulation may help shift Perpetually Plugged-In youth toward healthier engagement styles, while trauma-informed supports may be more effective for Burned-Out Browsers.

Third, platforms and policymakers should consider how design features and moderation practices interact with vulnerable offline contexts, particularly for youth facing adversity.

#### Limitations and future directions

This study has several limitations. First, data were cross-sectional, limiting our ability to establish temporal ordering or observe within-person transitions between engagement styles over time. Although we used causal discovery and Bayesian network–based interventions to estimate counterfactual effects, these inferences still rely on modeling assumptions, such as no substantial unmeasured confounding, correct variable discretization, and adequate representation of the true causal structure, and should be interpreted as hypothesis-generating rather than definitive causal estimates.

Second, all measures, including social media experiences, device use intensity, adversity, bullying, discrimination, and mental health outcomes, were self-reported and therefore subject to recall error, social desirability bias, and shared-method variance. The segmentation variables capture perceived experiences and motivations (e.g., comparison, identity exploration, emotional support) but do not directly measure platform-specific behaviors, content exposures, or algorithmic feed characteristics that may shape experiences in important ways.

Third, the segmentation solution reflects patterns in the specific set of items included in the survey. Different measures (e.g., distinguishing specific platforms, types of interaction such as posting versus passive scrolling, or differentiating online versus offline support-seeking) could yield alternative typologies. Relatedly, the five-cluster solution showed modest separation (silhouette width), suggesting meaningful but not sharply delineated boundaries between groups, and some youth may shift between profiles depending on circumstances.

Fourth, causal modeling required discretizing variables into a limited number of categories to ensure computational tractability. Discretization may reduce information, obscure nonlinearities, and affect estimated relationships. In addition, while we imposed constraints to prevent temporally implausible edges (e.g., downstream psychosocial variables causing demographics or childhood adversity), other directionality decisions remain uncertain in observational data, and alternative DAGs may fit similarly well.

Despite these limitations, combining population-representative segmentation with causal Bayesian network modeling offers a rigorous framework for moving beyond screen-time narratives and generating actionable hypotheses about intervention-relevant mechanisms linking social media engagement styles to youth wellbeing.

## Conclusion

This study demonstrates that youth social media engagement is best understood as part of a broader system of emotional regulation, social context, and structural vulnerability. By integrating segmentation with model-based causal discovery, we show that similar patterns of distressing online experiences can arise from distinct pathways and carry different implications for intervention. Recognizing this heterogeneity is essential for designing more effective, equitable strategies that support youth wellbeing in an increasingly digital world.

## Methods

### Study Design and Sample

Data were drawn from the 2024 Youth Mental Health Tracker (YMHT) survey, a cross-sectional, nationally representative survey of 4,509 individuals aged 10-24 years residing in the United States. The survey was administered from April through May 2024 using two complementary recruitment strategies. The first was a probability-based sample fielded by NORC at the University of Chicago through its AmeriSpeak panel, supplemented with an opt-in sample from the Prodege panel and calibrated using NORC’s proprietary calibration technique (NORC, 2025). Surveys were available in English and Spanish and could be completed online or via telephone. The second was a nonprobability sample recruited through targeted social media outreach to target harder-to-reach youth groups. All analyses for this manuscript were completed on the NORC Amerispeak with Prodege supplement sample to ensure population representativeness and because the primary analytic aim was not subgroup-specific, making the larger, targeted social media sample unnecessary for this purpose. Analyses were also restricted to participants aged 15-24 years (N = 2,563), as youth in this age range completed the longer, more comprehensive version of the survey. The study protocol and sampling strategy were reviewed and approved by the YMHT’s Subject Matter Expert Advisory Board and Youth Advisory Board to ensure ethical and age-appropriate implementation. Ethical approval was obtained from both the NORC Institutional Review Board (24-02-1653) and the Advarra IRB (Pro00076919). To promote participant safety, items on suicidal ideation or distress were immediately followed by crisis resource information (988 Suicide & Crisis Lifeline, Crisis Text Line). All survey responses were de-identified.

### Survey Instrument and Content

Survey development followed a mixed evidence- and youth-engagement process. A comprehensive literature review and focus groups with the Subject Matter Expert and Youth Advisory Boards informed item selection. Items were drawn from validated instruments and cognitively tested with youth to ensure comprehension and relevance across diverse populations.

Domains included: (1) demographics and socioeconomic context; (2) mental wellbeing and health perceptions; (3) youth-driven mental health strategies and supports; (4) risk and protective factors; (5) negative life experiences; and (6) access to and utilization of mental health care (Supplementary 5). Measures comprised a combination of items drawn from validated instruments, single-item adaptations capturing core constructs, and study-specific items developed and cognitively tested for this survey. Where full validated scales were not used, items were selected or designed to capture key dimensions of youth mental health and social context while minimizing respondent burden.

### Data Analysis

Data preparation and analysis were performed in R. Analyses focused on identifying distinct subgroups of youth based on a set of social media and digital use variables representing affective, behavioral, and identity-related dimensions of youth social media engagement (Table 2). Because the segmentation inputs include both social media specific items and broader device intensity/self-regulation items, we interpret the resulting clusters as “social media engagement styles embedded within broader digital routines,” rather than purely platform-specific behaviors. Analyses were limited to respondents with complete data on these variables and valid survey weights. Observations with missing data were excluded (2.7% of the total NORC sample).

#### Segmentation

To capture the multidimensional nature of social media engagement, we used Gower distance to compute pairwise dissimilarities among respondents. Gower distance was selected because it accommodates heterogeneous variable types within a single similarity metric while preserving the interpretability of each item’s contribution to overall dissimilarity. Binary variables contributed a value of 0 or 1 depending on concordance. To identify the optimal number of latent segments, we used both silhouette width analysis and the elbow method using total weighted sum of distances to medoids (the clustering objective function). Average silhouette widths were computed for cluster solutions ranging from 2 to 8 clusters. The optimal number of clusters was determined based on the maximum average silhouette width and the point of inflection (“elbow”) in the within-cluster sum of squares plot, indicating diminishing returns in explanatory gain. We then applied a weighted k-medoids clustering algorithm (partitioning around medoids; PAM) to identify the final segments. Respondent-level sampling weights, provided by NORC to ensure national representativeness via their TrueNorth calibration procedure aligning AmeriSpeak and supplementary opt-in panel data (NORC, 2021), were incorporated into the clustering. Based on a dissimilarity matrix, the algorithm minimizes the weighted sum of distances between each observation and the medoid of its assigned cluster, where weights reflect the relative contribution of each observation. The medoid is defined as the observation within a cluster that minimizes the total distance to all other observations in that cluster. Segment labels were developed iteratively by the research team to capture the most descriptive and distinctive behavioral and attitudinal characteristics of each cluster, based on comparative profile patterns. Cluster assignments were appended to the respondent-level dataset for subsequent causal analysis.

#### Causal Bayesian Network Modeling

We used a causal Bayesian network (BN) to model interdependencies among social media engagement segment membership, mental health, wellbeing, social context, adverse experiences and demographic characteristics. The directed acyclic graph (DAG) structure was learned directly from the data using causal discovery, which infers conditional dependency relationships by combining statistical tests of independence with model-based scoring. This approach enables identification of both direct and indirect causal pathways while accounting for observed confounding within the dataset.

We employed a previously validated hybrid causal discovery procedure (Butcher et al., 2021) that combines constraint-based independence testing to inform the initial graph structure with a score-based Markov Chain Monte Carlo (OrderMCMC) optimization over node orderings, using the Quotient Normalized Maximum Likelihood (qNML) criterion as the scoring function. This hybrid approach balances computational efficiency with improved structural and interventional accuracy and has been shown to outperform constraint-only methods in synthetic benchmark evaluations, particularly in moderate- to high-dimensional settings. Due to the computational complexity of unconstrained structure learning, model estimation was performed on a high-performance cloud computing instance.

### Variable selection and preprocessing

We selected 34 variables spanning demographics, mental health, wellbeing, adverse experiences, social context, and social media engagement (Table 1), guided by theoretical relevance and interpretability as distinct system components. All variables were categorical or ordinal; continuous measures were discretized into 2-5 levels to balance information retention with computational tractability and to ensure robust structure learning given the sample size.

To improve plausibility and reduce the search space, we incorporated domain-informed constraints that precluded logically impossible or temporally implausible relationships. Constraints enforced that demographic characteristics (age, gender, race/ethnicity, LGBTQ identity), adverse childhood experiences and discrimination could not be caused by psychosocial or behavioral variables, and social media engagement segment membership. These constraints ensured that the learned graph respected basic temporal ordering and substantive theory while preserving flexibility for data-driven discovery.

Structural performance was assessed using established graph-based metrics, including V-structure precision and recall, which indicated strong recovery of conditional dependency patterns. We simulated hypothetical interventions by fixing the exposure node to alternative levels in the Bayesian network and estimating resulting outcome probabilities using likelihood-weighted sampling. Effects were reported as odds ratios (ORs) contrasting the probability of the target outcome under exposed versus unexposed conditions, optionally conditional on additional evidence.

## Supporting information

All supplementary materials

## Funding

This study was funded by Pivotal Philanthropic Foundation.

## Acknowledgements

No acknowledgements.

## Disclosure

Authors are employees and/or owners of Surgo Health, a Public Benefit Corporation.

## Data Availability Statement

All data produced in the present study are available upon reasonable request to the authors.

## Notes

### Author Declarations

NORC Institutional Review Board (24-02-1653) and Advarra IRB (Pro00076919) gave ethical approval for this work

