## Supplementary material for "Heterogeneity in Youth Social Media Engagement and Its Pathways to Mental Health and Wellbeing": All supplementary materials

### Supplementary 1: Segmentation assessment metrics

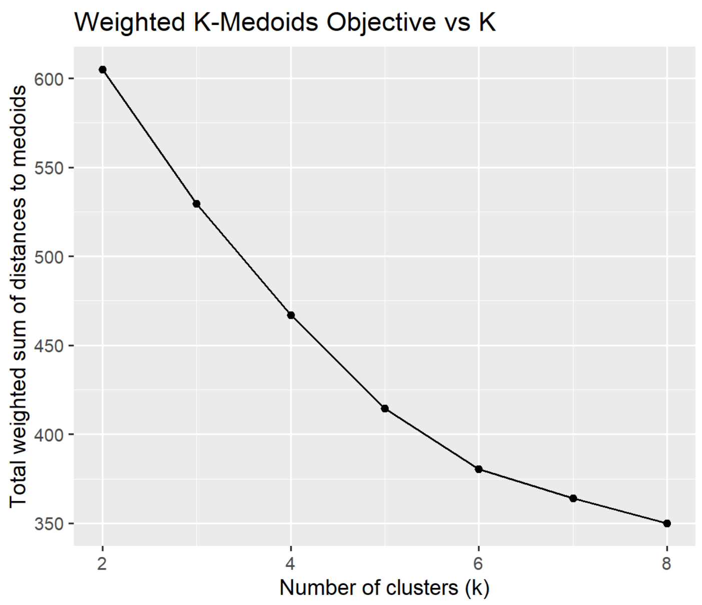


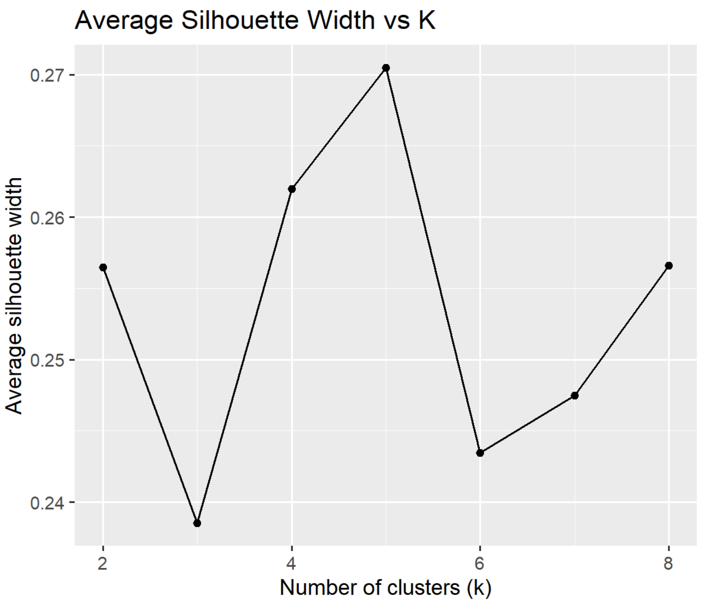

Supplementary 2: Full DAG

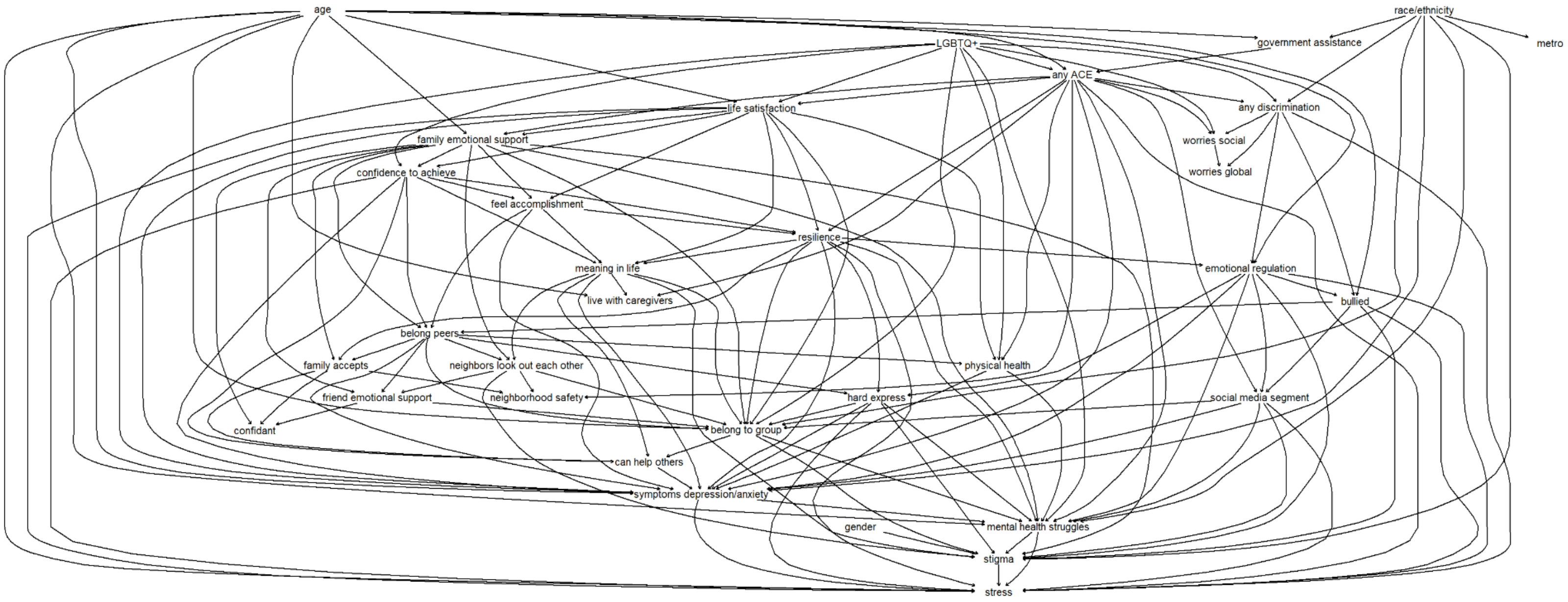

### Supplementary 3: Counterfactual interventions on upstream factors to segment

*Positive Engagers as the reference segment*

| intervention | Target segment | OR | Lower CI | Upper CI | sig |
|---|---|---|---|---|---|
| bullied:Disagree->Agree | Burned-Out Browsers | 0.67 | 0.55 | 0.82 | TRUE |
| government assistance:Assistance->No Assistance | Burned-Out Browsers | 0.79 | 0.65 | 0.97 | TRUE |
| hard express:Disagree->Agree | Burned-Out Browsers | 0.95 | 0.78 | 1.16 | FALSE |
| friend emotional support:Agree->Disagree | Burned-Out Browsers | 0.96 | 0.78 | 1.17 | FALSE |
| physical health:Good->Poor | Burned-Out Browsers | 0.97 | 0.80 | 1.19 | FALSE |
| stress:Disagree->Agree | Burned-Out Browsers | 0.97 | 0.80 | 1.19 | FALSE |
| gender:Woman->Another | Burned-Out Browsers | 0.97 | 0.80 | 1.19 | FALSE |
| belong peers:Agree->Disagree | Burned-Out Browsers | 0.98 | 0.80 | 1.20 | FALSE |
| metro:Metro Area->Non-Metro Area | Burned-Out Browsers | 0.99 | 0.81 | 1.20 | FALSE |
| live with caregivers:No->Yes | Burned-Out Browsers | 0.99 | 0.81 | 1.20 | FALSE |
| confidant:Agree->Disagree | Burned-Out Browsers | 0.99 | 0.81 | 1.21 | FALSE |
| any discrimination:0->1 | Burned-Out Browsers | 0.99 | 0.81 | 1.21 | FALSE |
| meaning in life:Agree->Disagree | Burned-Out Browsers | 0.99 | 0.81 | 1.21 | FALSE |
| race/ethnicity:white->asian | Burned-Out Browsers | 0.99 | 0.82 | 1.21 | FALSE |
| neighbors look out each other:Agree->Disagree | Burned-Out Browsers | 1.00 | 0.82 | 1.22 | FALSE |
| race/ethnicity:white->black | Burned-Out Browsers | 1.00 | 0.82 | 1.22 | FALSE |
| family accepts:Agree->Disagree | Burned-Out Browsers | 1.01 | 0.82 | 1.23 | FALSE |
| worries social:0->1 | Burned-Out Browsers | 1.01 | 0.82 | 1.23 | FALSE |
| worries global:0->1 | Burned-Out Browsers | 1.01 | 0.82 | 1.23 | FALSE |
| race/ethnicity:white->other | Burned-Out Browsers | 1.01 | 0.83 | 1.23 | FALSE |

| | | | | | |
|---|---|---|---|---|---|
| gender:Woman->Man | Burned-Out Browsers | 1.01 | 0.83 | 1.23 | FALSE |
| mental health struggles:no_minimal_mh_struggles->mh_struggles_daily_impact | Burned-Out Browsers | 1.01 | 0.83 | 1.23 | FALSE |
| symptoms depression/anxiety:No->Yes | Burned-Out Browsers | 1.01 | 0.83 | 1.24 | FALSE |
| belong to group:Agree->Disagree | Burned-Out Browsers | 1.03 | 0.84 | 1.25 | FALSE |
| stigma:Disagree->Agree | Burned-Out Browsers | 1.03 | 0.84 | 1.25 | FALSE |
| can help others:Agree->Disagree | Burned-Out Browsers | 1.03 | 0.85 | 1.26 | FALSE |
| family emotional support:Agree->Disagree | Burned-Out Browsers | 1.03 | 0.85 | 1.26 | FALSE |
| neighborhood safety:Agree->Disagree | Burned-Out Browsers | 1.04 | 0.85 | 1.26 | FALSE |
| race/ethnicity:white->hispanic | Burned-Out Browsers | 1.06 | 0.87 | 1.29 | FALSE |
| feel accomplishment:Agree->Disagree | Burned-Out Browsers | 1.08 | 0.88 | 1.31 | FALSE |
| confidence to achieve:Agree->Disagree | Burned-Out Browsers | 1.12 | 0.92 | 1.37 | FALSE |
| life satisfaction:Satisfied->Dissatisfied | Burned-Out Browsers | 1.13 | 0.93 | 1.39 | FALSE |
| resilience:Agree->Disagree | Burned-Out Browsers | 1.30 | 1.07 | 1.59 | TRUE |
| LGBTQ+:Not LGBTQ->LGBTQ | Burned-Out Browsers | 1.36 | 1.11 | 1.66 | TRUE |
| age:15-17->18-24 | Burned-Out Browsers | 1.49 | 1.22 | 1.81 | TRUE |
| emotional regulation:Disagree->Agree | Burned-Out Browsers | 2.30 | 1.88 | 2.81 | TRUE |
| any ACE:0->1 | Burned-Out Browsers | 3.63 | 2.94 | 4.47 | TRUE |
| bullied:Disagree->Agree | Light touch users | 0.50 | 0.39 | 0.63 | TRUE |
| any discrimination:0->1 | Light touch users | 0.89 | 0.70 | 1.12 | FALSE |
| metro:Metro Area->Non-Metro Area | Light touch users | 0.94 | 0.75 | 1.19 | FALSE |
| emotional regulation:Disagree->Agree | Light touch users | 0.95 | 0.75 | 1.20 | FALSE |
| friend emotional support:Agree->Disagree | Light touch users | 0.95 | 0.75 | 1.20 | FALSE |
| meaning in life:Agree->Disagree | Light touch users | 0.95 | 0.75 | 1.20 | FALSE |

| | | | | | |
|---|---|---|---|---|---|
| can help others:Agree->Disagree | Light touch users | 0.95 | 0.76 | 1.20 | FALSE |
| symptoms depression/anxiety:No->Yes | Light touch users | 0.96 | 0.76 | 1.21 | FALSE |
| live with caregivers:No->Yes | Light touch users | 0.96 | 0.77 | 1.21 | FALSE |
| neighborhood safety:Agree->Disagree | Light touch users | 0.97 | 0.77 | 1.22 | FALSE |
| belong peers:Agree->Disagree | Light touch users | 0.97 | 0.77 | 1.22 | FALSE |
| stigma:Disagree->Agree | Light touch users | 0.98 | 0.78 | 1.24 | FALSE |
| feel accomplishment:Agree->Disagree | Light touch users | 0.98 | 0.78 | 1.23 | FALSE |
| race/ethnicity:white->asian | Light touch users | 0.99 | 0.78 | 1.24 | FALSE |
| government assistance:Assistance->No Assistance | Light touch users | 0.99 | 0.79 | 1.25 | FALSE |
| worries social:0->1 | Light touch users | 1.00 | 0.79 | 1.25 | FALSE |
| race/ethnicity:white->black | Light touch users | 1.00 | 0.79 | 1.26 | FALSE |
| hard express:Disagree->Agree | Light touch users | 1.00 | 0.79 | 1.26 | FALSE |
| LGBTQ+:Not LGBTQ->LGBTQ | Light touch users | 1.00 | 0.79 | 1.27 | FALSE |
| race/ethnicity:white->hispanic | Light touch users | 1.00 | 0.80 | 1.26 | FALSE |
| life satisfaction:Satisfied->Dissatisfied | Light touch users | 1.00 | 0.80 | 1.27 | FALSE |
| stress:Disagree->Agree | Light touch users | 1.01 | 0.80 | 1.27 | FALSE |
| race/ethnicity:white->other | Light touch users | 1.01 | 0.80 | 1.27 | FALSE |
| mental health struggles:no_minimal_mh_struggles->mh_struggles_daily_impact | Light touch users | 1.01 | 0.80 | 1.28 | FALSE |
| confidence to achieve:Agree->Disagree | Light touch users | 1.02 | 0.80 | 1.28 | FALSE |
| gender:Woman->Man | Light touch users | 1.02 | 0.81 | 1.29 | FALSE |
| resilience:Agree->Disagree | Light touch users | 1.02 | 0.81 | 1.29 | FALSE |
| family emotional support:Agree->Disagree | Light touch users | 1.03 | 0.81 | 1.30 | FALSE |
| age:15-17->18-24 | Light touch users | 1.03 | 0.82 | 1.29 | FALSE |

| | | | | | |
|---|---|---|---|---|---|
| neighbors look out each other:Agree->Disagree | Light touch users | 1.03 | 0.82 | 1.30 | FALSE |
| belong to group:Agree->Disagree | Light touch users | 1.04 | 0.82 | 1.30 | FALSE |
| gender:Woman->Another | Light touch users | 1.06 | 0.84 | 1.34 | FALSE |
| physical health:Good->Poor | Light touch users | 1.06 | 0.84 | 1.34 | FALSE |
| worries global:0->1 | Light touch users | 1.06 | 0.84 | 1.34 | FALSE |
| family accepts:Agree->Disagree | Light touch users | 1.07 | 0.85 | 1.35 | FALSE |
| confidant:Agree->Disagree | Light touch users | 1.07 | 0.85 | 1.35 | FALSE |
| any ACE:0->1 | Light touch users | 1.19 | 0.94 | 1.51 | FALSE |
| government assistance:Assistance->No Assistance | Perpetually Plugged-In | 0.87 | 0.72 | 1.05 | FALSE |
| meaning in life:Agree->Disagree | Perpetually Plugged-In | 0.95 | 0.79 | 1.14 | FALSE |
| metro:Metro Area->Non-Metro Area | Perpetually Plugged-In | 0.97 | 0.81 | 1.17 | FALSE |
| gender:Woman->Another | Perpetually Plugged-In | 0.98 | 0.81 | 1.18 | FALSE |
| family emotional support:Agree->Disagree | Perpetually Plugged-In | 0.98 | 0.81 | 1.18 | FALSE |
| stress:Disagree->Agree | Perpetually Plugged-In | 0.98 | 0.82 | 1.18 | FALSE |
| gender:Woman->Man | Perpetually Plugged-In | 0.99 | 0.82 | 1.19 | FALSE |
| stigma:Disagree->Agree | Perpetually Plugged-In | 0.99 | 0.82 | 1.19 | FALSE |
| confidant:Agree->Disagree | Perpetually Plugged-In | 0.99 | 0.82 | 1.19 | FALSE |
| friend emotional support:Agree->Disagree | Perpetually Plugged-In | 0.99 | 0.82 | 1.19 | FALSE |
| neighborhood safety:Agree->Disagree | Perpetually Plugged-In | 1.00 | 0.83 | 1.20 | FALSE |
| hard express:Disagree->Agree | Perpetually Plugged-In | 1.00 | 0.83 | 1.21 | FALSE |
| feel accomplishment:Agree->Disagree | Perpetually Plugged-In | 1.01 | 0.83 | 1.21 | FALSE |
| worries global:0->1 | Perpetually Plugged-In | 1.01 | 0.84 | 1.21 | FALSE |
| belong to group:Agree->Disagree | Perpetually Plugged-In | 1.01 | 0.84 | 1.21 | FALSE |
| live with caregivers:No->Yes | Perpetually Plugged-In | 1.01 | 0.84 | 1.21 | FALSE |

| | | | | | |
|---|---|---|---|---|---|
| symptoms depression/anxiety:No->Yes | Perpetually Plugged-In | 1.01 | 0.84 | 1.22 | FALSE |
| neighbors look out each other:Agree->Disagree | Perpetually Plugged-In | 1.01 | 0.84 | 1.22 | FALSE |
| belong peers:Agree->Disagree | Perpetually Plugged-In | 1.02 | 0.84 | 1.22 | FALSE |
| physical health:Good->Poor | Perpetually Plugged-In | 1.02 | 0.84 | 1.22 | FALSE |
| can help others:Agree->Disagree | Perpetually Plugged-In | 1.02 | 0.85 | 1.23 | FALSE |
| race/ethnicity:white->asian | Perpetually Plugged-In | 1.03 | 0.86 | 1.24 | FALSE |
| family accepts:Agree->Disagree | Perpetually Plugged-In | 1.03 | 0.86 | 1.24 | FALSE |
| mental health struggles:no_minimal_mh_struggles->mh_struggles_daily_impact | Perpetually Plugged-In | 1.04 | 0.86 | 1.25 | FALSE |
| worries social:0->1 | Perpetually Plugged-In | 1.04 | 0.87 | 1.25 | FALSE |
| race/ethnicity:white->black | Perpetually Plugged-In | 1.05 | 0.87 | 1.26 | FALSE |
| race/ethnicity:white->other | Perpetually Plugged-In | 1.05 | 0.87 | 1.26 | FALSE |
| race/ethnicity:white->hispanic | Perpetually Plugged-In | 1.06 | 0.88 | 1.27 | FALSE |
| life satisfaction:Satisfied->Dissatisfied | Perpetually Plugged-In | 1.11 | 0.93 | 1.34 | FALSE |
| confidence to achieve:Agree->Disagree | Perpetually Plugged-In | 1.12 | 0.93 | 1.35 | FALSE |
| any discrimination:0->1 | Perpetually Plugged-In | 1.26 | 1.04 | 1.51 | TRUE |
| resilience:Agree->Disagree | Perpetually Plugged-In | 1.34 | 1.11 | 1.61 | TRUE |
| bullied:Disagree->Agree | Perpetually Plugged-In | 1.35 | 1.12 | 1.61 | TRUE |
| age:15-17->18-24 | Perpetually Plugged-In | 1.36 | 1.13 | 1.63 | TRUE |
| LGBTQ+:Not LGBTQ->LGBTQ | Perpetually Plugged-In | 1.37 | 1.13 | 1.65 | TRUE |
| any ACE:0->1 | Perpetually Plugged-In | 2.79 | 2.31 | 3.38 | TRUE |
| emotional regulation:Disagree->Agree | Perpetually Plugged-In | 3.29 | 2.72 | 3.98 | TRUE |
| bullied:Disagree->Agree | Practical navigators | 0.42 | 0.34 | 0.51 | TRUE |
| any discrimination:0->1 | Practical navigators | 0.87 | 0.71 | 1.06 | FALSE |
| government assistance:Assistance->No Assistance | Practical navigators | 0.89 | 0.73 | 1.09 | FALSE |

| | | | | | |
|---|---|---|---|---|---|
| stigma:Disagree->Agree | Practical navigators | 0.95 | 0.78 | 1.17 | FALSE |
| friend emotional support:Agree->Disagree | Practical navigators | 0.95 | 0.78 | 1.16 | FALSE |
| race/ethnicity:white->black | Practical navigators | 0.96 | 0.78 | 1.17 | FALSE |
| gender:Woman->Another | Practical navigators | 0.96 | 0.79 | 1.18 | FALSE |
| worries social:0->1 | Practical navigators | 0.97 | 0.79 | 1.19 | FALSE |
| feel accomplishment:Agree->Disagree | Practical navigators | 0.97 | 0.79 | 1.19 | FALSE |
| race/ethnicity:white->other | Practical navigators | 0.97 | 0.79 | 1.19 | FALSE |
| neighborhood safety:Agree->Disagree | Practical navigators | 0.98 | 0.80 | 1.20 | FALSE |
| confidence to achieve:Agree->Disagree | Practical navigators | 0.98 | 0.80 | 1.20 | FALSE |
| race/ethnicity:white->asian | Practical navigators | 0.98 | 0.80 | 1.21 | FALSE |
| meaning in life:Agree->Disagree | Practical navigators | 0.99 | 0.80 | 1.21 | FALSE |
| neighbors look out each other:Agree->Disagree | Practical navigators | 0.99 | 0.81 | 1.21 | FALSE |
| stress:Disagree->Agree | Practical navigators | 1.00 | 0.81 | 1.22 | FALSE |
| worries global:0->1 | Practical navigators | 1.00 | 0.82 | 1.23 | FALSE |
| live with caregivers:No->Yes | Practical navigators | 1.00 | 0.82 | 1.23 | FALSE |
| emotional regulation:Disagree->Agree | Practical navigators | 1.00 | 0.82 | 1.23 | FALSE |
| belong to group:Agree->Disagree | Practical navigators | 1.00 | 0.82 | 1.23 | FALSE |
| belong peers:Agree->Disagree | Practical navigators | 1.01 | 0.82 | 1.23 | FALSE |
| resilience:Agree->Disagree | Practical navigators | 1.01 | 0.82 | 1.24 | FALSE |
| metro:Metro Area->Non-Metro Area | Practical navigators | 1.01 | 0.82 | 1.23 | FALSE |
| life satisfaction:Satisfied->Dissatisfied | Practical navigators | 1.01 | 0.82 | 1.24 | FALSE |
| can help others:Agree->Disagree | Practical navigators | 1.01 | 0.82 | 1.24 | FALSE |
| mental health struggles:no_minimal_mh_struggles->mh_struggles_daily_impact | Practical navigators | 1.01 | 0.83 | 1.24 | FALSE |

| | | | | | |
|---|---|---|---|---|---|
| family emotional support:Agree->Disagree | Practical navigators | 1.01 | 0.83 | 1.24 | FALSE |
| physical health:Good->Poor | Practical navigators | 1.01 | 0.83 | 1.24 | FALSE |
| race/ethnicity:white->hispanic | Practical navigators | 1.01 | 0.83 | 1.24 | FALSE |
| hard express:Disagree->Agree | Practical navigators | 1.04 | 0.85 | 1.27 | FALSE |
| symptoms depression/anxiety:No->Yes | Practical navigators | 1.04 | 0.85 | 1.27 | FALSE |
| gender:Woman->Man | Practical navigators | 1.05 | 0.85 | 1.28 | FALSE |
| confidant:Agree->Disagree | Practical navigators | 1.06 | 0.86 | 1.29 | FALSE |
| family accepts:Agree->Disagree | Practical navigators | 1.06 | 0.86 | 1.30 | FALSE |
| LGBTQ+:Not LGBTQ->LGBTQ | Practical navigators | 1.14 | 0.93 | 1.41 | FALSE |
| age:15-17->18-24 | Practical navigators | 1.23 | 1.00 | 1.51 | TRUE |
| any ACE:0->1 | Practical navigators | 1.89 | 1.54 | 2.33 | TRUE |

*Burned Out Browsers as the reference segment*

| intervention | Target segment | OR | Lower CI | Upper CI | sig |
|---|---|---|---|---|---|
| any ACE:0->1 | Light touch users | 0.35 | 0.28 | 0.44 | TRUE |
| emotional regulation:Disagree->Agree | Light touch users | 0.39 | 0.31 | 0.48 | TRUE |
| age:15-17->18-24 | Light touch users | 0.72 | 0.58 | 0.89 | TRUE |
| resilience:Agree->Disagree | Light touch users | 0.75 | 0.60 | 0.93 | TRUE |
| LGBTQ+:Not LGBTQ->LGBTQ | Light touch users | 0.77 | 0.63 | 0.96 | TRUE |
| bullied:Disagree->Agree | Light touch users | 0.79 | 0.63 | 0.99 | TRUE |

| | | | | | |
|---|---|---|---|---|---|
| any discrimination:0->1 | Light touch users | 0.89 | 0.72 | 1.10 | FALSE |
| confidence to achieve:Agree->Disagree | Light touch users | 0.91 | 0.73 | 1.13 | FALSE |
| race/ethnicity:white->black | Light touch users | 0.91 | 0.74 | 1.13 | FALSE |
| life satisfaction:Satisfied->Dissatisfied | Light touch users | 0.92 | 0.74 | 1.14 | FALSE |
| feel accomplishment:Agree->Disagree | Light touch users | 0.94 | 0.76 | 1.17 | FALSE |
| metro:Metro Area->Non-Metro Area | Light touch users | 0.95 | 0.77 | 1.17 | FALSE |
| stigma:Disagree->Agree | Light touch users | 0.95 | 0.77 | 1.18 | FALSE |
| family emotional support:Agree->Disagree | Light touch users | 0.96 | 0.77 | 1.18 | FALSE |
| race/ethnicity:white->other | Light touch users | 0.97 | 0.78 | 1.20 | FALSE |
| worries social:0->1 | Light touch users | 0.97 | 0.78 | 1.20 | FALSE |
| family accepts:Agree->Disagree | Light touch users | 0.97 | 0.79 | 1.20 | FALSE |
| stress:Disagree->Agree | Light touch users | 0.97 | 0.79 | 1.21 | FALSE |
| friend emotional support:Agree->Disagree | Light touch users | 0.97 | 0.79 | 1.20 | FALSE |
| physical health:Good->Poor | Light touch users | 0.98 | 0.79 | 1.21 | FALSE |
| belong peers:Agree->Disagree | Light touch users | 0.98 | 0.79 | 1.22 | FALSE |
| gender:Woman->Another | Light touch users | 0.98 | 0.79 | 1.22 | FALSE |

| | | | | | |
|---|---|---|---|---|---|
| symptoms depression/anxiety:No->Yes | Light touch users | 0.98 | 0.80 | 1.22 | FALSE |
| race/ethnicity:white->hispanic | Light touch users | 0.99 | 0.80 | 1.22 | FALSE |
| mental health struggles:no_minimal_mh_struggles->mh_struggles_daily_impact | Light touch users | 1.00 | 0.81 | 1.24 | FALSE |
| race/ethnicity:white->asian | Light touch users | 1.01 | 0.82 | 1.24 | FALSE |
| worries global:0->1 | Light touch users | 1.01 | 0.82 | 1.25 | FALSE |
| neighbors look out each other:Agree->Disagree | Light touch users | 1.02 | 0.82 | 1.26 | FALSE |
| gender:Woman->Man | Light touch users | 1.02 | 0.82 | 1.26 | FALSE |
| can help others:Agree->Disagree | Light touch users | 1.02 | 0.82 | 1.26 | FALSE |
| confidant:Agree->Disagree | Light touch users | 1.02 | 0.83 | 1.26 | FALSE |
| hard express:Disagree->Agree | Light touch users | 1.03 | 0.83 | 1.28 | FALSE |
| neighborhood safety:Agree->Disagree | Light touch users | 1.03 | 0.84 | 1.28 | FALSE |
| live with caregivers:No->Yes | Light touch users | 1.04 | 0.84 | 1.28 | FALSE |
| belong to group:Agree->Disagree | Light touch users | 1.04 | 0.84 | 1.28 | FALSE |
| meaning in life:Agree->Disagree | Light touch users | 1.04 | 0.84 | 1.29 | FALSE |
| government assistance:Assistance->No Assistance | Light touch users | 1.17 | 0.95 | 1.45 | FALSE |
| any ACE:0->1 | Perpetually Plugged-In | 0.84 | 0.71 | 0.99 | TRUE |

| | | | | | |
|---|---|---|---|---|---|
| age:15-17->18-24 | Perpetually Plugged-In | 0.88 | 0.75 | 1.04 | FALSE |
| stigma:Disagree->Agree | Perpetually Plugged-In | 0.96 | 0.82 | 1.13 | FALSE |
| worries social:0->1 | Perpetually Plugged-In | 0.97 | 0.83 | 1.14 | FALSE |
| live with caregivers:No->Yes | Perpetually Plugged-In | 0.97 | 0.83 | 1.14 | FALSE |
| family emotional support:Agree->Disagree | Perpetually Plugged-In | 0.98 | 0.83 | 1.14 | FALSE |
| can help others:Agree->Disagree | Perpetually Plugged-In | 0.99 | 0.84 | 1.16 | FALSE |
| belong peers:Agree->Disagree | Perpetually Plugged-In | 0.99 | 0.84 | 1.16 | FALSE |
| confidence to achieve:Agree->Disagree | Perpetually Plugged-In | 1.00 | 0.85 | 1.17 | FALSE |
| LGBTQ+:Not LGBTQ->LGBTQ | Perpetually Plugged-In | 1.00 | 0.85 | 1.17 | FALSE |
| friend emotional support:Agree->Disagree | Perpetually Plugged-In | 1.00 | 0.85 | 1.17 | FALSE |
| hard express:Disagree->Agree | Perpetually Plugged-In | 1.00 | 0.85 | 1.18 | FALSE |
| physical health:Good->Poor | Perpetually Plugged-In | 1.00 | 0.85 | 1.18 | FALSE |
| gender:Woman->Another | Perpetually Plugged-In | 1.00 | 0.86 | 1.18 | FALSE |
| feel accomplishment:Agree->Disagree | Perpetually Plugged-In | 1.01 | 0.86 | 1.18 | FALSE |
| symptoms depression/anxiety:No->Yes | Perpetually Plugged-In | 1.01 | 0.86 | 1.18 | FALSE |
| worries global:0->1 | Perpetually Plugged-In | 1.01 | 0.86 | 1.18 | FALSE |

| | | | | | |
|---|---|---|---|---|---|
| neighbors look out each other:Agree->Disagree | Perpetually Plugged-In | 1.01 | 0.86 | 1.18 | FALSE |
| stress:Disagree->Agree | Perpetually Plugged-In | 1.01 | 0.86 | 1.19 | FALSE |
| neighborhood safety:Agree->Disagree | Perpetually Plugged-In | 1.01 | 0.86 | 1.19 | FALSE |
| government assistance:Assistance->No Assistance | Perpetually Plugged-In | 1.02 | 0.87 | 1.19 | FALSE |
| belong to group:Agree->Disagree | Perpetually Plugged-In | 1.02 | 0.87 | 1.19 | FALSE |
| meaning in life:Agree->Disagree | Perpetually Plugged-In | 1.02 | 0.87 | 1.20 | FALSE |
| family accepts:Agree->Disagree | Perpetually Plugged-In | 1.02 | 0.87 | 1.20 | FALSE |
| life satisfaction:Satisfied->Dissatisfied | Perpetually Plugged-In | 1.02 | 0.87 | 1.20 | FALSE |
| gender:Woman->Man | Perpetually Plugged-In | 1.03 | 0.87 | 1.20 | FALSE |
| race/ethnicity:white->other | Perpetually Plugged-In | 1.03 | 0.87 | 1.21 | FALSE |
| race/ethnicity:white->hispanic | Perpetually Plugged-In | 1.03 | 0.88 | 1.21 | FALSE |
| confidant:Agree->Disagree | Perpetually Plugged-In | 1.03 | 0.88 | 1.21 | FALSE |
| mental health struggles:no_minimal_mh_struggles->mh_struggles_daily_impact | Perpetually Plugged-In | 1.03 | 0.88 | 1.21 | FALSE |
| metro:Metro Area->Non-Metro Area | Perpetually Plugged-In | 1.03 | 0.88 | 1.21 | FALSE |
| race/ethnicity:white->black | Perpetually Plugged-In | 1.05 | 0.90 | 1.24 | FALSE |
| race/ethnicity:white->asian | Perpetually Plugged-In | 1.06 | 0.90 | 1.25 | FALSE |

| | | | | | |
|---|---|---|---|---|---|
| resilience:Agree->Disagree | Perpetually Plugged-In | 1.07 | 0.91 | 1.25 | FALSE |
| any discrimination:0->1 | Perpetually Plugged-In | 1.19 | 1.02 | 1.40 | TRUE |
| emotional regulation:Disagree->Agree | Perpetually Plugged-In | 1.42 | 1.20 | 1.68 | TRUE |
| bullied:Disagree->Agree | Perpetually Plugged-In | 1.97 | 1.67 | 2.31 | TRUE |
| any ACE:0->1 | Positive engagers | 0.31 | 0.25 | 0.38 | TRUE |
| emotional regulation:Disagree->Agree | Positive engagers | 0.44 | 0.36 | 0.54 | TRUE |
| age:15-17->18-24 | Positive engagers | 0.64 | 0.53 | 0.79 | TRUE |
| LGBTQ+:Not LGBTQ->LGBTQ | Positive engagers | 0.74 | 0.61 | 0.91 | TRUE |
| resilience:Agree->Disagree | Positive engagers | 0.78 | 0.63 | 0.95 | TRUE |
| life satisfaction:Satisfied->Dissatisfied | Positive engagers | 0.90 | 0.74 | 1.10 | FALSE |
| any discrimination:0->1 | Positive engagers | 0.96 | 0.79 | 1.17 | FALSE |
| race/ethnicity:white->black | Positive engagers | 0.97 | 0.79 | 1.18 | FALSE |
| meaning in life:Agree->Disagree | Positive engagers | 0.97 | 0.80 | 1.19 | FALSE |
| gender:Woman->Man | Positive engagers | 0.97 | 0.80 | 1.19 | FALSE |
| can help others:Agree->Disagree | Positive engagers | 0.97 | 0.80 | 1.19 | FALSE |
| race/ethnicity:white->other | Positive engagers | 0.98 | 0.80 | 1.19 | FALSE |
| confidence to achieve:Agree->Disagree | Positive engagers | 0.98 | 0.80 | 1.19 | FALSE |
| friend emotional support:Agree->Disagree | Positive engagers | 0.98 | 0.80 | 1.20 | FALSE |
| feel accomplishment:Agree->Disagree | Positive engagers | 0.98 | 0.80 | 1.20 | FALSE |
| neighborhood safety:Agree->Disagree | Positive engagers | 0.99 | 0.81 | 1.20 | FALSE |

| | | | | | |
|---|---|---|---|---|---|
| belong peers:Agree->Disagree | Positive engagers | 0.99 | 0.81 | 1.21 | FALSE |
| belong to group:Agree->Disagree | Positive engagers | 1.00 | 0.82 | 1.22 | FALSE |
| metro:Metro Area->Non-Metro Area | Positive engagers | 1.00 | 0.82 | 1.22 | FALSE |
| worries global:0->1 | Positive engagers | 1.00 | 0.82 | 1.22 | FALSE |
| race/ethnicity:white->hispanic | Positive engagers | 1.01 | 0.82 | 1.23 | FALSE |
| hard express:Disagree->Agree | Positive engagers | 1.01 | 0.82 | 1.23 | FALSE |
| neighbors look out each other:Agree->Disagree | Positive engagers | 1.01 | 0.83 | 1.23 | FALSE |
| family accepts:Agree->Disagree | Positive engagers | 1.01 | 0.83 | 1.23 | FALSE |
| stigma:Disagree->Agree | Positive engagers | 1.02 | 0.83 | 1.24 | FALSE |
| family emotional support:Agree->Disagree | Positive engagers | 1.02 | 0.84 | 1.24 | FALSE |
| physical health:Good->Poor | Positive engagers | 1.02 | 0.84 | 1.25 | FALSE |
| symptoms depression/anxiety:No->Yes | Positive engagers | 1.03 | 0.84 | 1.25 | FALSE |
| confidant:Agree->Disagree | Positive engagers | 1.03 | 0.84 | 1.25 | FALSE |
| race/ethnicity:white->asian | Positive engagers | 1.03 | 0.84 | 1.26 | FALSE |
| mental health struggles:no_minimal_mh_struggles->mh_struggles_daily_impact | Positive engagers | 1.04 | 0.86 | 1.27 | FALSE |
| live with caregivers:No->Yes | Positive engagers | 1.05 | 0.86 | 1.28 | FALSE |
| stress:Disagree->Agree | Positive engagers | 1.05 | 0.86 | 1.28 | FALSE |
| worries social:0->1 | Positive engagers | 1.05 | 0.86 | 1.29 | FALSE |
| gender:Woman->Another | Positive engagers | 1.07 | 0.88 | 1.31 | FALSE |
| government assistance:Assistance->No Assistance | Positive engagers | 1.22 | 1.00 | 1.49 | FALSE |
| bullied:Disagree->Agree | Positive engagers | 1.60 | 1.31 | 1.96 | TRUE |
| emotional regulation:Disagree->Agree | Practical navigators | 0.45 | 0.37 | 0.54 | TRUE |

| | | | | | |
|---|---|---|---|---|---|
| any ACE:0->1 | Practical navigators | 0.58 | 0.48 | 0.70 | TRUE |
| bullied:Disagree->Agree | Practical navigators | 0.60 | 0.49 | 0.72 | TRUE |
| resilience:Agree->Disagree | Practical navigators | 0.79 | 0.66 | 0.95 | TRUE |
| age:15-17->18-24 | Practical navigators | 0.80 | 0.66 | 0.96 | TRUE |
| any discrimination:0->1 | Practical navigators | 0.80 | 0.66 | 0.96 | TRUE |
| LGBTQ+:Not LGBTQ->LGBTQ | Practical navigators | 0.83 | 0.69 | 0.99 | TRUE |
| race/ethnicity:white->black | Practical navigators | 0.92 | 0.77 | 1.10 | FALSE |
| life satisfaction:Satisfied->Dissatisfied | Practical navigators | 0.94 | 0.78 | 1.12 | FALSE |
| confidence to achieve:Agree->Disagree | Practical navigators | 0.95 | 0.80 | 1.14 | FALSE |
| race/ethnicity:white->hispanic | Practical navigators | 0.96 | 0.80 | 1.15 | FALSE |
| meaning in life:Agree->Disagree | Practical navigators | 0.96 | 0.81 | 1.15 | FALSE |
| neighborhood safety:Agree->Disagree | Practical navigators | 0.97 | 0.81 | 1.16 | FALSE |
| gender:Woman->Another | Practical navigators | 0.97 | 0.81 | 1.17 | FALSE |
| race/ethnicity:white->other | Practical navigators | 0.97 | 0.81 | 1.17 | FALSE |
| belong peers:Agree->Disagree | Practical navigators | 0.97 | 0.81 | 1.17 | FALSE |
| family emotional support:Agree->Disagree | Practical navigators | 0.98 | 0.81 | 1.17 | FALSE |
| stigma:Disagree->Agree | Practical navigators | 0.98 | 0.82 | 1.17 | FALSE |
| feel accomplishment:Agree->Disagree | Practical navigators | 0.99 | 0.82 | 1.18 | FALSE |
| neighbors look out each other:Agree->Disagree | Practical navigators | 1.00 | 0.83 | 1.20 | FALSE |

| | | | | | |
|---|---|---|---|---|---|
| confidant:Agree->Disagree | Practical navigators | 1.00 | 0.83 | 1.20 | FALSE |
| worries social:0->1 | Practical navigators | 1.00 | 0.83 | 1.20 | FALSE |
| race/ethnicity:white->asian | Practical navigators | 1.00 | 0.83 | 1.20 | FALSE |
| can help others:Agree->Disagree | Practical navigators | 1.00 | 0.84 | 1.20 | FALSE |
| hard express:Disagree->Agree | Practical navigators | 1.01 | 0.84 | 1.21 | FALSE |
| metro:Metro Area->Non-Metro Area | Practical navigators | 1.01 | 0.84 | 1.21 | FALSE |
| physical health:Good->Poor | Practical navigators | 1.02 | 0.85 | 1.22 | FALSE |
| worries global:0->1 | Practical navigators | 1.02 | 0.85 | 1.22 | FALSE |
| belong to group:Agree->Disagree | Practical navigators | 1.02 | 0.85 | 1.22 | FALSE |
| stress:Disagree->Agree | Practical navigators | 1.02 | 0.85 | 1.22 | FALSE |
| gender:Woman->Man | Practical navigators | 1.02 | 0.85 | 1.22 | FALSE |
| live with caregivers:No->Yes | Practical navigators | 1.03 | 0.86 | 1.23 | FALSE |
| friend emotional support:Agree->Disagree | Practical navigators | 1.03 | 0.86 | 1.23 | FALSE |
| family accepts:Agree->Disagree | Practical navigators | 1.04 | 0.87 | 1.24 | FALSE |
| symptoms depression/anxiety:No->Yes | Practical navigators | 1.04 | 0.87 | 1.24 | FALSE |
| mental health struggles:no_minimal_mh_struggles->mh_struggles_daily_impact | Practical navigators | 1.04 | 0.87 | 1.24 | FALSE |

| | | | | | |
|---|---|---|---|---|---|
| government assistance:Assistance->No Assistance | Practical navigators | 1.10 | 0.91 | 1.31 | FALSE |

*Perpetually Plugged-In as the reference segment*

| intervention | Target segment | OR | Lower CI | Upper CI | sig |
|---|---|---|---|---|---|
| bullied:Disagree->Agree | Burned-Out Browsers | 0.51 | 0.43 | 0.60 | TRUE |
| emotional regulation:Disagree->Agree | Burned-Out Browsers | 0.68 | 0.58 | 0.81 | TRUE |
| any discrimination:0->1 | Burned-Out Browsers | 0.83 | 0.71 | 0.98 | TRUE |
| race/ethnicity:white->other | Burned-Out Browsers | 0.93 | 0.80 | 1.10 | FALSE |
| government assistance:Assistance->No Assistance | Burned-Out Browsers | 0.96 | 0.81 | 1.12 | FALSE |
| metro:Metro Area->Non-Metro Area | Burned-Out Browsers | 0.96 | 0.82 | 1.13 | FALSE |
| meaning in life:Agree->Disagree | Burned-Out Browsers | 0.96 | 0.82 | 1.13 | FALSE |
| symptoms depression/anxiety:No->Yes | Burned-Out Browsers | 0.97 | 0.82 | 1.13 | FALSE |
| resilience:Agree->Disagree | Burned-Out Browsers | 0.97 | 0.83 | 1.13 | FALSE |
| can help others:Agree->Disagree | Burned-Out Browsers | 0.97 | 0.82 | 1.14 | FALSE |
| confidence to achieve:Agree->Disagree | Burned-Out Browsers | 0.97 | 0.83 | 1.14 | FALSE |
| life satisfaction:Satisfied->Dissatisfied | Burned-Out Browsers | 0.98 | 0.84 | 1.15 | FALSE |

| | | | | | |
|---|---|---|---|---|---|
| worries social:0->1 | Burned-Out Browsers | 0.98 | 0.84 | 1.15 | FALSE |
| race/ethnicity:white->black | Burned-Out Browsers | 0.98 | 0.84 | 1.16 | FALSE |
| confidant:Agree->Disagree | Burned-Out Browsers | 0.99 | 0.84 | 1.16 | FALSE |
| LGBTQ+:Not LGBTQ->LGBTQ | Burned-Out Browsers | 0.99 | 0.84 | 1.16 | FALSE |
| race/ethnicity:white->hispanic | Burned-Out Browsers | 0.99 | 0.84 | 1.16 | FALSE |
| neighborhood safety:Agree->Disagree | Burned-Out Browsers | 0.99 | 0.84 | 1.16 | FALSE |
| race/ethnicity:white->asian | Burned-Out Browsers | 0.99 | 0.84 | 1.16 | FALSE |
| physical health:Good->Poor | Burned-Out Browsers | 0.99 | 0.84 | 1.16 | FALSE |
| family emotional support:Agree->Disagree | Burned-Out Browsers | 0.99 | 0.84 | 1.16 | FALSE |
| family accepts:Agree->Disagree | Burned-Out Browsers | 1.00 | 0.85 | 1.17 | FALSE |
| gender:Woman->Man | Burned-Out Browsers | 1.00 | 0.85 | 1.17 | FALSE |
| belong peers:Agree->Disagree | Burned-Out Browsers | 1.00 | 0.85 | 1.17 | FALSE |
| hard express:Disagree->Agree | Burned-Out Browsers | 1.00 | 0.85 | 1.17 | FALSE |
| friend emotional support:Agree->Disagree | Burned-Out Browsers | 1.00 | 0.86 | 1.18 | FALSE |
| neighbors look out each other:Agree->Disagree | Burned-Out Browsers | 1.01 | 0.86 | 1.18 | FALSE |
| stigma:Disagree->Agree | Burned-Out Browsers | 1.01 | 0.86 | 1.18 | FALSE |

| | | | | | |
|---|---|---|---|---|---|
| feel accomplishment:Agree->Disagree | Burned-Out Browsers | 1.01 | 0.86 | 1.19 | FALSE |
| gender:Woman->Another | Burned-Out Browsers | 1.01 | 0.86 | 1.19 | FALSE |
| worries global:0->1 | Burned-Out Browsers | 1.02 | 0.87 | 1.20 | FALSE |
| mental health struggles:no_minimal_mh_struggles->mh_struggles_daily_impact | Burned-Out Browsers | 1.03 | 0.87 | 1.20 | FALSE |
| belong to group:Agree->Disagree | Burned-Out Browsers | 1.03 | 0.87 | 1.21 | FALSE |
| stress:Disagree->Agree | Burned-Out Browsers | 1.03 | 0.88 | 1.21 | FALSE |
| live with caregivers:No->Yes | Burned-Out Browsers | 1.04 | 0.89 | 1.22 | FALSE |
| age:15-17->18-24 | Burned-Out Browsers | 1.08 | 0.92 | 1.27 | FALSE |
| any ACE:0->1 | Burned-Out Browsers | 1.19 | 1.01 | 1.40 | TRUE |
| emotional regulation:Disagree->Agree | Light touch users | 0.27 | 0.22 | 0.33 | TRUE |
| bullied:Disagree->Agree | Light touch users | 0.40 | 0.32 | 0.49 | TRUE |
| any ACE:0->1 | Light touch users | 0.41 | 0.33 | 0.50 | TRUE |
| any discrimination:0->1 | Light touch users | 0.69 | 0.57 | 0.84 | TRUE |
| resilience:Agree->Disagree | Light touch users | 0.71 | 0.58 | 0.87 | TRUE |
| LGBTQ+:Not LGBTQ->LGBTQ | Light touch users | 0.73 | 0.60 | 0.90 | TRUE |
| age:15-17->18-24 | Light touch users | 0.76 | 0.62 | 0.93 | TRUE |

| | | | | | |
|---|---|---|---|---|---|
| confidence to achieve:Agree->Disagree | Light touch users | 0.87 | 0.71 | 1.06 | FALSE |
| life satisfaction:Satisfied->Dissatisfied | Light touch users | 0.87 | 0.71 | 1.06 | FALSE |
| feel accomplishment:Agree->Disagree | Light touch users | 0.88 | 0.72 | 1.08 | FALSE |
| race/ethnicity:white->other | Light touch users | 0.91 | 0.74 | 1.11 | FALSE |
| can help others:Agree->Disagree | Light touch users | 0.93 | 0.76 | 1.14 | FALSE |
| gender:Woman->Another | Light touch users | 0.95 | 0.78 | 1.16 | FALSE |
| neighbors look out each other:Agree->Disagree | Light touch users | 0.96 | 0.79 | 1.18 | FALSE |
| race/ethnicity:white->black | Light touch users | 0.97 | 0.79 | 1.18 | FALSE |
| stress:Disagree->Agree | Light touch users | 0.97 | 0.80 | 1.18 | FALSE |
| confidant:Agree->Disagree | Light touch users | 0.97 | 0.79 | 1.19 | FALSE |
| race/ethnicity:white->asian | Light touch users | 0.98 | 0.80 | 1.19 | FALSE |
| worries global:0->1 | Light touch users | 0.98 | 0.80 | 1.19 | FALSE |
| hard express:Disagree->Agree | Light touch users | 0.98 | 0.80 | 1.20 | FALSE |
| gender:Woman->Man | Light touch users | 0.98 | 0.80 | 1.19 | FALSE |
| belong peers:Agree->Disagree | Light touch users | 0.98 | 0.81 | 1.20 | FALSE |
| live with caregivers:No->Yes | Light touch users | 0.98 | 0.81 | 1.20 | FALSE |

| | | | | | |
|---|---|---|---|---|---|
| race/ethnicity:white->hispanic | Light touch users | 0.99 | 0.81 | 1.20 | FALSE |
| family emotional support:Agree->Disagree | Light touch users | 0.99 | 0.81 | 1.21 | FALSE |
| worries social:0->1 | Light touch users | 0.99 | 0.81 | 1.21 | FALSE |
| neighborhood safety:Agree->Disagree | Light touch users | 1.00 | 0.81 | 1.22 | FALSE |
| friend emotional support:Agree->Disagree | Light touch users | 1.00 | 0.82 | 1.22 | FALSE |
| mental health struggles:no_minimal_mh_struggles->mh_struggles_daily_impact | Light touch users | 1.00 | 0.82 | 1.23 | FALSE |
| metro:Metro Area->Non-Metro Area | Light touch users | 1.01 | 0.83 | 1.23 | FALSE |
| family accepts:Agree->Disagree | Light touch users | 1.02 | 0.83 | 1.24 | FALSE |
| symptoms depression/anxiety:No->Yes | Light touch users | 1.02 | 0.83 | 1.24 | FALSE |
| physical health:Good->Poor | Light touch users | 1.02 | 0.84 | 1.25 | FALSE |
| belong to group:Agree->Disagree | Light touch users | 1.03 | 0.84 | 1.25 | FALSE |
| stigma:Disagree->Agree | Light touch users | 1.03 | 0.84 | 1.25 | FALSE |
| meaning in life:Agree->Disagree | Light touch users | 1.03 | 0.85 | 1.26 | FALSE |
| government assistance:Assistance->No Assistance | Light touch users | 1.16 | 0.95 | 1.42 | FALSE |
| emotional regulation:Disagree->Agree | Positive engagers | 0.30 | 0.25 | 0.36 | TRUE |

| | | | | | |
|---|---|---|---|---|---|
| any ACE:0->1 | Positive engagers | 0.37 | 0.30 | 0.44 | TRUE |
| age:15-17->18-24 | Positive engagers | 0.69 | 0.58 | 0.84 | TRUE |
| LGBTQ+:Not LGBTQ->LGBTQ | Positive engagers | 0.71 | 0.59 | 0.86 | TRUE |
| resilience:Agree->Disagree | Positive engagers | 0.73 | 0.61 | 0.88 | TRUE |
| bullied:Disagree->Agree | Positive engagers | 0.76 | 0.64 | 0.91 | TRUE |
| any discrimination:0->1 | Positive engagers | 0.82 | 0.69 | 0.99 | TRUE |
| life satisfaction:Satisfied->Dissatisfied | Positive engagers | 0.88 | 0.73 | 1.06 | FALSE |
| family emotional support:Agree->Disagree | Positive engagers | 0.89 | 0.74 | 1.07 | FALSE |
| race/ethnicity:white->black | Positive engagers | 0.92 | 0.77 | 1.11 | FALSE |
| race/ethnicity:white->other | Positive engagers | 0.93 | 0.77 | 1.12 | FALSE |
| confidence to achieve:Agree->Disagree | Positive engagers | 0.93 | 0.77 | 1.12 | FALSE |
| feel accomplishment:Agree->Disagree | Positive engagers | 0.93 | 0.77 | 1.12 | FALSE |
| race/ethnicity:white->hispanic | Positive engagers | 0.95 | 0.79 | 1.14 | FALSE |
| neighbors look out each other:Agree->Disagree | Positive engagers | 0.95 | 0.79 | 1.15 | FALSE |
| physical health:Good->Poor | Positive engagers | 0.97 | 0.80 | 1.16 | FALSE |
| stress:Disagree->Agree | Positive engagers | 0.97 | 0.81 | 1.16 | FALSE |
| worries global:0->1 | Positive engagers | 0.97 | 0.80 | 1.17 | FALSE |
| race/ethnicity:white->asian | Positive engagers | 0.97 | 0.81 | 1.17 | FALSE |
| belong peers:Agree->Disagree | Positive engagers | 0.97 | 0.81 | 1.17 | FALSE |
| meaning in life:Agree->Disagree | Positive engagers | 0.97 | 0.81 | 1.17 | FALSE |
| confidant:Agree->Disagree | Positive engagers | 0.98 | 0.81 | 1.17 | FALSE |
| gender:Woman->Man | Positive engagers | 0.98 | 0.81 | 1.18 | FALSE |
| metro:Metro Area->Non-Metro Area | Positive engagers | 0.98 | 0.81 | 1.18 | FALSE |

| | | | | | |
|---|---|---|---|---|---|
| friend emotional support:Agree->Disagree | Positive engagers | 0.99 | 0.82 | 1.19 | FALSE |
| live with caregivers:No->Yes | Positive engagers | 0.99 | 0.83 | 1.20 | FALSE |
| worries social:0->1 | Positive engagers | 1.01 | 0.84 | 1.21 | FALSE |
| mental health struggles:no_minimal_mh_struggles->mh_struggles_daily_impact | Positive engagers | 1.01 | 0.84 | 1.21 | FALSE |
| family accepts:Agree->Disagree | Positive engagers | 1.02 | 0.85 | 1.23 | FALSE |
| neighborhood safety:Agree->Disagree | Positive engagers | 1.02 | 0.85 | 1.23 | FALSE |
| gender:Woman->Another | Positive engagers | 1.03 | 0.85 | 1.24 | FALSE |
| belong to group:Agree->Disagree | Positive engagers | 1.03 | 0.85 | 1.24 | FALSE |
| hard express:Disagree->Agree | Positive engagers | 1.03 | 0.86 | 1.24 | FALSE |
| symptoms depression/anxiety:No->Yes | Positive engagers | 1.05 | 0.87 | 1.26 | FALSE |
| can help others:Agree->Disagree | Positive engagers | 1.05 | 0.87 | 1.27 | FALSE |
| stigma:Disagree->Agree | Positive engagers | 1.06 | 0.88 | 1.27 | FALSE |
| government assistance:Assistance->No Assistance | Positive engagers | 1.22 | 1.02 | 1.48 | TRUE |
| emotional regulation:Disagree->Agree | Practical navigators | 0.31 | 0.26 | 0.36 | TRUE |
| bullied:Disagree->Agree | Practical navigators | 0.31 | 0.26 | 0.38 | TRUE |
| any ACE:0->1 | Practical navigators | 0.67 | 0.57 | 0.80 | TRUE |
| any discrimination:0->1 | Practical navigators | 0.70 | 0.60 | 0.83 | TRUE |
| resilience:Agree->Disagree | Practical navigators | 0.74 | 0.63 | 0.88 | TRUE |
| LGBTQ+:Not LGBTQ->LGBTQ | Practical navigators | 0.82 | 0.70 | 0.97 | TRUE |

| | | | | | |
|---|---|---|---|---|---|
| life satisfaction:Satisfied->Dissatisfied | Practical navigators | 0.90 | 0.77 | 1.06 | FALSE |
| race/ethnicity:white->hispanic | Practical navigators | 0.92 | 0.78 | 1.08 | FALSE |
| age:15-17->18-24 | Practical navigators | 0.92 | 0.78 | 1.09 | FALSE |
| confidence to achieve:Agree->Disagree | Practical navigators | 0.93 | 0.78 | 1.09 | FALSE |
| feel accomplishment:Agree->Disagree | Practical navigators | 0.93 | 0.79 | 1.10 | FALSE |
| race/ethnicity:white->black | Practical navigators | 0.94 | 0.80 | 1.11 | FALSE |
| race/ethnicity:white->asian | Practical navigators | 0.94 | 0.80 | 1.12 | FALSE |
| family emotional support:Agree->Disagree | Practical navigators | 0.95 | 0.81 | 1.12 | FALSE |
| friend emotional support:Agree->Disagree | Practical navigators | 0.96 | 0.81 | 1.13 | FALSE |
| race/ethnicity:white->other | Practical navigators | 0.96 | 0.82 | 1.14 | FALSE |
| mental health struggles:no_minimal_mh_struggles->mh_struggles_daily_impact | Practical navigators | 0.97 | 0.82 | 1.15 | FALSE |
| belong peers:Agree->Disagree | Practical navigators | 0.98 | 0.83 | 1.15 | FALSE |
| belong to group:Agree->Disagree | Practical navigators | 0.98 | 0.83 | 1.15 | FALSE |
| metro:Metro Area->Non-Metro Area | Practical navigators | 0.98 | 0.83 | 1.16 | FALSE |
| stigma:Disagree->Agree | Practical navigators | 0.99 | 0.84 | 1.17 | FALSE |
| symptoms depression/anxiety:No->Yes | Practical navigators | 0.99 | 0.84 | 1.17 | FALSE |

| | | | | | |
|---|---|---|---|---|---|
| neighbors look out each other:Agree->Disagree | Practical navigators | 1.00 | 0.85 | 1.18 | FALSE |
| family accepts:Agree->Disagree | Practical navigators | 1.00 | 0.85 | 1.18 | FALSE |
| gender:Woman->Man | Practical navigators | 1.01 | 0.86 | 1.19 | FALSE |
| physical health:Good->Poor | Practical navigators | 1.01 | 0.86 | 1.20 | FALSE |
| stress:Disagree->Agree | Practical navigators | 1.01 | 0.86 | 1.20 | FALSE |
| can help others:Agree->Disagree | Practical navigators | 1.02 | 0.86 | 1.20 | FALSE |
| live with caregivers:No->Yes | Practical navigators | 1.02 | 0.87 | 1.21 | FALSE |
| gender:Woman->Another | Practical navigators | 1.03 | 0.87 | 1.21 | FALSE |
| confidant:Agree->Disagree | Practical navigators | 1.03 | 0.87 | 1.22 | FALSE |
| government assistance:Assistance->No Assistance | Practical navigators | 1.03 | 0.87 | 1.22 | FALSE |
| neighborhood safety:Agree->Disagree | Practical navigators | 1.04 | 0.88 | 1.22 | FALSE |
| worries social:0->1 | Practical navigators | 1.04 | 0.88 | 1.22 | FALSE |
| hard express:Disagree->Agree | Practical navigators | 1.05 | 0.89 | 1.24 | FALSE |
| meaning in life:Agree->Disagree | Practical navigators | 1.06 | 0.90 | 1.25 | FALSE |
| worries global:0->1 | Practical navigators | 1.07 | 0.91 | 1.26 | FALSE |

*Light Touch Users as the reference segment*

| intervention | target_segment | OR | lower CI | upper CI | sig |
|---|---|---|---|---|---|
| government assistance:Assistance->No Assistance | Burned-Out Browsers | 0.81 | 0.66 | 1.00 | FALSE |
| friend emotional support:Agree->Disagree | Burned-Out Browsers | 0.91 | 0.74 | 1.13 | FALSE |
| live with caregivers:No->Yes | Burned-Out Browsers | 0.93 | 0.76 | 1.16 | FALSE |
| mental health struggles:no_minimal_mh_struggles->mh_struggles_daily_impact | Burned-Out Browsers | 0.95 | 0.77 | 1.17 | FALSE |
| meaning in life:Agree->Disagree | Burned-Out Browsers | 0.95 | 0.77 | 1.18 | FALSE |
| can help others:Agree->Disagree | Burned-Out Browsers | 0.96 | 0.78 | 1.19 | FALSE |
| gender:Woman->Another | Burned-Out Browsers | 0.98 | 0.79 | 1.20 | FALSE |
| worries global:0->1 | Burned-Out Browsers | 0.98 | 0.80 | 1.22 | FALSE |
| neighbors look out each other:Agree->Disagree | Burned-Out Browsers | 0.99 | 0.80 | 1.22 | FALSE |
| belong peers:Agree->Disagree | Burned-Out Browsers | 0.99 | 0.80 | 1.23 | FALSE |
| gender:Woman->Man | Burned-Out Browsers | 0.99 | 0.80 | 1.23 | FALSE |
| stigma:Disagree->Agree | Burned-Out Browsers | 1.00 | 0.81 | 1.23 | FALSE |
| worries social:0->1 | Burned-Out Browsers | 1.00 | 0.81 | 1.23 | FALSE |
| symptoms depression/anxiety:No->Yes | Burned-Out Browsers | 1.01 | 0.82 | 1.25 | FALSE |

| | | | | | |
|---|---|---|---|---|---|
| family accepts:Agree->Disagree | Burned-Out Browsers | 1.01 | 0.82 | 1.25 | FALSE |
| stress:Disagree->Agree | Burned-Out Browsers | 1.02 | 0.82 | 1.25 | FALSE |
| neighborhood safety:Agree->Disagree | Burned-Out Browsers | 1.02 | 0.83 | 1.26 | FALSE |
| belong to group:Agree->Disagree | Burned-Out Browsers | 1.02 | 0.83 | 1.26 | FALSE |
| physical health:Good->Poor | Burned-Out Browsers | 1.03 | 0.83 | 1.27 | FALSE |
| life satisfaction:Satisfied->Dissatisfied | Burned-Out Browsers | 1.03 | 0.83 | 1.27 | FALSE |
| confidant:Agree->Disagree | Burned-Out Browsers | 1.03 | 0.83 | 1.28 | FALSE |
| metro:Metro Area->Non-Metro Area | Burned-Out Browsers | 1.04 | 0.84 | 1.28 | FALSE |
| race/ethnicity:white->asian | Burned-Out Browsers | 1.04 | 0.84 | 1.29 | FALSE |
| hard express:Disagree->Agree | Burned-Out Browsers | 1.05 | 0.85 | 1.30 | FALSE |
| feel accomplishment:Agree->Disagree | Burned-Out Browsers | 1.06 | 0.86 | 1.31 | FALSE |
| family emotional support:Agree->Disagree | Burned-Out Browsers | 1.07 | 0.86 | 1.32 | FALSE |
| race/ethnicity:white->other | Burned-Out Browsers | 1.07 | 0.87 | 1.33 | FALSE |
| race/ethnicity:white->hispanic | Burned-Out Browsers | 1.07 | 0.87 | 1.33 | FALSE |
| confidence to achieve:Agree->Disagree | Burned-Out Browsers | 1.08 | 0.87 | 1.33 | FALSE |
| race/ethnicity:white->black | Burned-Out Browsers | 1.09 | 0.88 | 1.35 | FALSE |

| | | | | | |
|---|---|---|---|---|---|
| any discrimination:0->1 | Burned-Out Browsers | 1.20 | 0.97 | 1.48 | FALSE |
| bullied:Disagree->Agree | Burned-Out Browsers | 1.24 | 0.99 | 1.55 | FALSE |
| LGBTQ+:Not LGBTQ->LGBTQ | Burned-Out Browsers | 1.27 | 1.02 | 1.57 | TRUE |
| resilience:Agree->Disagree | Burned-Out Browsers | 1.34 | 1.08 | 1.67 | TRUE |
| age:15-17->18-24 | Burned-Out Browsers | 1.41 | 1.14 | 1.74 | TRUE |
| emotional regulation:Disagree->Agree | Burned-Out Browsers | 2.50 | 2.02 | 3.11 | TRUE |
| any ACE:0->1 | Burned-Out Browsers | 2.73 | 2.20 | 3.39 | TRUE |
| government assistance:Assistance->No Assistance | Perpetually Plugged-In | 0.92 | 0.75 | 1.13 | FALSE |
| meaning in life:Agree->Disagree | Perpetually Plugged-In | 0.93 | 0.76 | 1.13 | FALSE |
| mental health struggles:no_minimal_mh_struggles->mh_struggles_daily_impact | Perpetually Plugged-In | 0.94 | 0.77 | 1.15 | FALSE |
| worries global:0->1 | Perpetually Plugged-In | 0.97 | 0.79 | 1.18 | FALSE |
| stigma:Disagree->Agree | Perpetually Plugged-In | 0.98 | 0.80 | 1.20 | FALSE |
| confidant:Agree->Disagree | Perpetually Plugged-In | 0.99 | 0.81 | 1.20 | FALSE |
| belong peers:Agree->Disagree | Perpetually Plugged-In | 1.00 | 0.82 | 1.22 | FALSE |
| neighborhood safety:Agree->Disagree | Perpetually Plugged-In | 1.01 | 0.82 | 1.23 | FALSE |
| gender:Woman->Man | Perpetually Plugged-In | 1.01 | 0.83 | 1.23 | FALSE |

| | | | | | |
|---|---|---|---|---|---|
| worries social:0->1 | Perpetually Plugged-In | 1.01 | 0.83 | 1.23 | FALSE |
| race/ethnicity:white->asian | Perpetually Plugged-In | 1.01 | 0.83 | 1.24 | FALSE |
| physical health:Good->Poor | Perpetually Plugged-In | 1.01 | 0.83 | 1.24 | FALSE |
| family accepts:Agree->Disagree | Perpetually Plugged-In | 1.02 | 0.83 | 1.24 | FALSE |
| neighbors look out each other:Agree->Disagree | Perpetually Plugged-In | 1.02 | 0.83 | 1.24 | FALSE |
| symptoms depression/anxiety:No->Yes | Perpetually Plugged-In | 1.02 | 0.84 | 1.24 | FALSE |
| friend emotional support:Agree->Disagree | Perpetually Plugged-In | 1.02 | 0.84 | 1.25 | FALSE |
| race/ethnicity:white->hispanic | Perpetually Plugged-In | 1.03 | 0.84 | 1.25 | FALSE |
| family emotional support:Agree->Disagree | Perpetually Plugged-In | 1.03 | 0.84 | 1.25 | FALSE |
| metro:Metro Area->Non-Metro Area | Perpetually Plugged-In | 1.03 | 0.85 | 1.26 | FALSE |
| belong to group:Agree->Disagree | Perpetually Plugged-In | 1.03 | 0.84 | 1.26 | FALSE |
| gender:Woman->Another | Perpetually Plugged-In | 1.03 | 0.85 | 1.26 | FALSE |
| live with caregivers:No->Yes | Perpetually Plugged-In | 1.03 | 0.85 | 1.26 | FALSE |
| stress:Disagree->Agree | Perpetually Plugged-In | 1.04 | 0.85 | 1.27 | FALSE |
| can help others:Agree->Disagree | Perpetually Plugged-In | 1.04 | 0.86 | 1.27 | FALSE |
| feel accomplishment:Agree->Disagree | Perpetually Plugged-In | 1.05 | 0.86 | 1.28 | FALSE |

| | | | | | |
|---|---|---|---|---|---|
| race/ethnicity:white->other | Perpetually Plugged-In | 1.05 | 0.86 | 1.29 | FALSE |
| hard express:Disagree->Agree | Perpetually Plugged-In | 1.07 | 0.88 | 1.31 | FALSE |
| race/ethnicity:white->black | Perpetually Plugged-In | 1.09 | 0.90 | 1.33 | FALSE |
| confidence to achieve:Agree->Disagree | Perpetually Plugged-In | 1.12 | 0.91 | 1.37 | FALSE |
| life satisfaction:Satisfied->Dissatisfied | Perpetually Plugged-In | 1.13 | 0.92 | 1.37 | FALSE |
| LGBTQ+:Not LGBTQ->LGBTQ | Perpetually Plugged-In | 1.33 | 1.09 | 1.63 | TRUE |
| age:15-17->18-24 | Perpetually Plugged-In | 1.36 | 1.11 | 1.66 | TRUE |
| resilience:Agree->Disagree | Perpetually Plugged-In | 1.38 | 1.13 | 1.68 | TRUE |
| any discrimination:0->1 | Perpetually Plugged-In | 1.39 | 1.14 | 1.70 | TRUE |
| any ACE:0->1 | Perpetually Plugged-In | 2.35 | 1.92 | 2.88 | TRUE |
| bullied:Disagree->Agree | Perpetually Plugged-In | 2.48 | 2.01 | 3.05 | TRUE |
| emotional regulation:Disagree->Agree | Perpetually Plugged-In | 3.66 | 2.98 | 4.50 | TRUE |
| any ACE:0->1 | Positive engagers | 0.84 | 0.67 | 1.07 | FALSE |
| age:15-17->18-24 | Positive engagers | 0.93 | 0.74 | 1.17 | FALSE |
| LGBTQ+:Not LGBTQ->LGBTQ | Positive engagers | 0.96 | 0.75 | 1.22 | FALSE |
| worries global:0->1 | Positive engagers | 0.96 | 0.76 | 1.21 | FALSE |
| metro:Metro Area->Non-Metro Area | Positive engagers | 0.97 | 0.77 | 1.22 | FALSE |
| stigma:Disagree->Agree | Positive engagers | 0.97 | 0.77 | 1.22 | FALSE |
| belong peers:Agree->Disagree | Positive engagers | 0.97 | 0.77 | 1.23 | FALSE |

| | | | | | |
|---|---|---|---|---|---|
| gender:Woman->Another | Positive engagers | 0.97 | 0.77 | 1.23 | FALSE |
| worries social:0->1 | Positive engagers | 0.98 | 0.78 | 1.23 | FALSE |
| family accepts:Agree->Disagree | Positive engagers | 0.98 | 0.78 | 1.23 | FALSE |
| race/ethnicity:white->other | Positive engagers | 0.98 | 0.78 | 1.24 | FALSE |
| friend emotional support:Agree->Disagree | Positive engagers | 0.98 | 0.78 | 1.24 | FALSE |
| family emotional support:Agree->Disagree | Positive engagers | 0.99 | 0.79 | 1.24 | FALSE |
| meaning in life:Agree->Disagree | Positive engagers | 1.00 | 0.79 | 1.26 | FALSE |
| live with caregivers:No->Yes | Positive engagers | 1.00 | 0.79 | 1.26 | FALSE |
| can help others:Agree->Disagree | Positive engagers | 1.00 | 0.79 | 1.26 | FALSE |
| stress:Disagree->Agree | Positive engagers | 1.00 | 0.80 | 1.27 | FALSE |
| belong to group:Agree->Disagree | Positive engagers | 1.00 | 0.80 | 1.27 | FALSE |
| government assistance:Assistance->No Assistance | Positive engagers | 1.01 | 0.80 | 1.27 | FALSE |
| race/ethnicity:white->asian | Positive engagers | 1.02 | 0.81 | 1.28 | FALSE |
| life satisfaction:Satisfied->Dissatisfied | Positive engagers | 1.02 | 0.81 | 1.29 | FALSE |
| confidant:Agree->Disagree | Positive engagers | 1.03 | 0.81 | 1.29 | FALSE |
| neighbors look out each other:Agree->Disagree | Positive engagers | 1.03 | 0.82 | 1.29 | FALSE |
| symptoms depression/anxiety:No->Yes | Positive engagers | 1.03 | 0.82 | 1.30 | FALSE |
| confidence to achieve:Agree->Disagree | Positive engagers | 1.03 | 0.82 | 1.30 | FALSE |
| hard express:Disagree->Agree | Positive engagers | 1.03 | 0.82 | 1.30 | FALSE |
| resilience:Agree->Disagree | Positive engagers | 1.03 | 0.82 | 1.31 | FALSE |
| gender:Woman->Man | Positive engagers | 1.04 | 0.82 | 1.31 | FALSE |
| physical health:Good->Poor | Positive engagers | 1.05 | 0.83 | 1.32 | FALSE |

| | | | | | |
|---|---|---|---|---|---|
| mental health struggles:no_minimal_mh_struggles->mh_struggles_daily_impact | Positive engagers | 1.05 | 0.83 | 1.32 | FALSE |
| neighborhood safety:Agree->Disagree | Positive engagers | 1.06 | 0.84 | 1.34 | FALSE |
| race/ethnicity:white->hispanic | Positive engagers | 1.06 | 0.84 | 1.34 | FALSE |
| feel accomplishment:Agree->Disagree | Positive engagers | 1.07 | 0.85 | 1.35 | FALSE |
| race/ethnicity:white->black | Positive engagers | 1.08 | 0.86 | 1.35 | FALSE |
| emotional regulation:Disagree->Agree | Positive engagers | 1.15 | 0.91 | 1.45 | FALSE |
| any discrimination:0->1 | Positive engagers | 1.15 | 0.92 | 1.45 | FALSE |
| bullied:Disagree->Agree | Positive engagers | 1.89 | 1.48 | 2.40 | TRUE |
| bullied:Disagree->Agree | Practical navigators | 0.82 | 0.65 | 1.05 | FALSE |
| government assistance:Assistance->No Assistance | Practical navigators | 0.92 | 0.74 | 1.14 | FALSE |
| race/ethnicity:white->asian | Practical navigators | 0.94 | 0.76 | 1.17 | FALSE |
| live with caregivers:No->Yes | Practical navigators | 0.96 | 0.77 | 1.19 | FALSE |
| belong peers:Agree->Disagree | Practical navigators | 0.96 | 0.78 | 1.20 | FALSE |
| any discrimination:0->1 | Practical navigators | 0.97 | 0.78 | 1.20 | FALSE |
| worries global:0->1 | Practical navigators | 0.97 | 0.78 | 1.20 | FALSE |
| stress:Disagree->Agree | Practical navigators | 0.97 | 0.78 | 1.20 | FALSE |
| confidant:Agree->Disagree | Practical navigators | 0.97 | 0.78 | 1.21 | FALSE |
| metro:Metro Area->Non-Metro Area | Practical navigators | 0.97 | 0.78 | 1.21 | FALSE |
| can help others:Agree->Disagree | Practical navigators | 0.99 | 0.80 | 1.23 | FALSE |

| | | | | | |
|---|---|---|---|---|---|
| race/ethnicity:white->other | Practical navigators | 0.99 | 0.80 | 1.23 | FALSE |
| physical health:Good->Poor | Practical navigators | 1.00 | 0.80 | 1.23 | FALSE |
| worries social:0->1 | Practical navigators | 1.00 | 0.80 | 1.24 | FALSE |
| meaning in life:Agree->Disagree | Practical navigators | 1.00 | 0.80 | 1.24 | FALSE |
| confidence to achieve:Agree->Disagree | Practical navigators | 1.00 | 0.80 | 1.24 | FALSE |
| neighbors look out each other:Agree->Disagree | Practical navigators | 1.00 | 0.81 | 1.24 | FALSE |
| gender:Woman->Another | Practical navigators | 1.00 | 0.81 | 1.25 | FALSE |
| gender:Woman->Man | Practical navigators | 1.00 | 0.81 | 1.25 | FALSE |
| belong to group:Agree->Disagree | Practical navigators | 1.01 | 0.81 | 1.25 | FALSE |
| race/ethnicity:white->hispanic | Practical navigators | 1.01 | 0.81 | 1.25 | FALSE |
| family accepts:Agree->Disagree | Practical navigators | 1.01 | 0.81 | 1.25 | FALSE |
| hard express:Disagree->Agree | Practical navigators | 1.02 | 0.82 | 1.26 | FALSE |
| family emotional support:Agree->Disagree | Practical navigators | 1.02 | 0.83 | 1.27 | FALSE |
| race/ethnicity:white->black | Practical navigators | 1.03 | 0.83 | 1.28 | FALSE |
| symptoms depression/anxiety:No->Yes | Practical navigators | 1.03 | 0.83 | 1.27 | FALSE |
| stigma:Disagree->Agree | Practical navigators | 1.03 | 0.83 | 1.28 | FALSE |

| | | | | | |
|---|---|---|---|---|---|
| friend emotional support:Agree->Disagree | Practical navigators | 1.03 | 0.83 | 1.28 | FALSE |
| feel accomplishment:Agree->Disagree | Practical navigators | 1.04 | 0.84 | 1.29 | FALSE |
| neighborhood safety:Agree->Disagree | Practical navigators | 1.04 | 0.84 | 1.29 | FALSE |
| mental health struggles:no_minimal_mh_struggles->mh_struggles_daily_impact | Practical navigators | 1.05 | 0.85 | 1.30 | FALSE |
| resilience:Agree->Disagree | Practical navigators | 1.06 | 0.85 | 1.32 | FALSE |
| life satisfaction:Satisfied->Dissatisfied | Practical navigators | 1.06 | 0.85 | 1.32 | FALSE |
| emotional regulation:Disagree->Agree | Practical navigators | 1.08 | 0.87 | 1.34 | FALSE |
| age:15-17->18-24 | Practical navigators | 1.09 | 0.88 | 1.35 | FALSE |
| LGBTQ+:Not LGBTQ->LGBTQ | Practical navigators | 1.13 | 0.91 | 1.41 | FALSE |
| any ACE:0->1 | Practical navigators | 1.61 | 1.30 | 2.00 | TRUE |

*Practical navigators as the reference segment*

| intervention | target_segment | OR | Lower CI | Upper CI | sig |
|---|---|---|---|---|---|
| government assistance:Assistance->No Assistance | Burned-Out Browsers | 0.90 | 0.75 | 1.07 | FALSE |
| race/ethnicity:white->asian | Burned-Out Browsers | 0.95 | 0.79 | 1.14 | FALSE |
| gender:Woman->Man | Burned-Out Browsers | 0.95 | 0.80 | 1.14 | FALSE |
| neighbors look out each other:Agree->Disagree | Burned-Out Browsers | 0.96 | 0.80 | 1.15 | FALSE |

| | | | | | |
|---|---|---|---|---|---|
| live with caregivers:No->Yes | Burned-Out Browsers | 0.96 | 0.80 | 1.15 | FALSE |
| belong peers:Agree->Disagree | Burned-Out Browsers | 0.97 | 0.81 | 1.16 | FALSE |
| hard express:Disagree->Agree | Burned-Out Browsers | 0.98 | 0.82 | 1.17 | FALSE |
| family accepts:Agree->Disagree | Burned-Out Browsers | 0.98 | 0.82 | 1.18 | FALSE |
| stress:Disagree->Agree | Burned-Out Browsers | 0.99 | 0.82 | 1.18 | FALSE |
| symptoms depression/anxiety:No->Yes | Burned-Out Browsers | 0.99 | 0.82 | 1.18 | FALSE |
| can help others:Agree->Disagree | Burned-Out Browsers | 0.99 | 0.83 | 1.19 | FALSE |
| neighborhood safety:Agree->Disagree | Burned-Out Browsers | 1.00 | 0.83 | 1.19 | FALSE |
| metro:Metro Area->Non-Metro Area | Burned-Out Browsers | 1.00 | 0.83 | 1.19 | FALSE |
| physical health:Good->Poor | Burned-Out Browsers | 1.00 | 0.84 | 1.20 | FALSE |
| belong to group:Agree->Disagree | Burned-Out Browsers | 1.01 | 0.84 | 1.21 | FALSE |
| worries global:0->1 | Burned-Out Browsers | 1.01 | 0.84 | 1.21 | FALSE |
| gender:Woman->Another | Burned-Out Browsers | 1.01 | 0.84 | 1.21 | FALSE |
| worries social:0->1 | Burned-Out Browsers | 1.01 | 0.84 | 1.21 | FALSE |
| family emotional support:Agree->Disagree | Burned-Out Browsers | 1.01 | 0.84 | 1.21 | FALSE |
| mental health struggles:no_minimal_mh_struggles->mh_struggles_daily_impact | Burned-Out Browsers | 1.01 | 0.84 | 1.21 | FALSE |

| | | | | | |
|---|---|---|---|---|---|
| race/ethnicity:white->hispanic | Burned-Out Browsers | 1.01 | 0.85 | 1.21 | FALSE |
| confidant:Agree->Disagree | Burned-Out Browsers | 1.02 | 0.85 | 1.22 | FALSE |
| friend emotional support:Agree->Disagree | Burned-Out Browsers | 1.03 | 0.86 | 1.23 | FALSE |
| stigma:Disagree->Agree | Burned-Out Browsers | 1.03 | 0.86 | 1.23 | FALSE |
| meaning in life:Agree->Disagree | Burned-Out Browsers | 1.03 | 0.86 | 1.24 | FALSE |
| race/ethnicity:white->other | Burned-Out Browsers | 1.04 | 0.87 | 1.24 | FALSE |
| feel accomplishment:Agree->Disagree | Burned-Out Browsers | 1.04 | 0.87 | 1.25 | FALSE |
| confidence to achieve:Agree->Disagree | Burned-Out Browsers | 1.09 | 0.91 | 1.31 | FALSE |
| race/ethnicity:white->black | Burned-Out Browsers | 1.09 | 0.91 | 1.31 | FALSE |
| life satisfaction:Satisfied->Dissatisfied | Burned-Out Browsers | 1.11 | 0.92 | 1.33 | FALSE |
| any discrimination:0->1 | Burned-Out Browsers | 1.17 | 0.98 | 1.41 | FALSE |
| LGBTQ+:Not LGBTQ->LGBTQ | Burned-Out Browsers | 1.21 | 1.01 | 1.45 | TRUE |
| resilience:Agree->Disagree | Burned-Out Browsers | 1.21 | 1.01 | 1.46 | TRUE |
| age:15-17->18-24 | Burned-Out Browsers | 1.27 | 1.06 | 1.52 | TRUE |
| bullied:Disagree->Agree | Burned-Out Browsers | 1.64 | 1.34 | 2.00 | TRUE |
| any ACE:0->1 | Burned-Out Browsers | 1.73 | 1.44 | 2.08 | TRUE |

| | | | | | |
|---|---|---|---|---|---|
| emotional regulation:Disagree->Agree | Burned-Out Browsers | 2.25 | 1.87 | 2.71 | TRUE |
| any ACE:0->1 | Light touch users | 0.60 | 0.48 | 0.74 | TRUE |
| emotional regulation:Disagree->Agree | Light touch users | 0.83 | 0.67 | 1.03 | FALSE |
| age:15-17->18-24 | Light touch users | 0.87 | 0.70 | 1.07 | FALSE |
| LGBTQ+:Not LGBTQ->LGBTQ | Light touch users | 0.90 | 0.72 | 1.12 | FALSE |
| life satisfaction:Satisfied->Dissatisfied | Light touch users | 0.92 | 0.74 | 1.15 | FALSE |
| race/ethnicity:white->black | Light touch users | 0.94 | 0.76 | 1.18 | FALSE |
| race/ethnicity:white->hispanic | Light touch users | 0.95 | 0.76 | 1.17 | FALSE |
| worries global:0->1 | Light touch users | 0.95 | 0.77 | 1.18 | FALSE |
| confidant:Agree->Disagree | Light touch users | 0.95 | 0.77 | 1.18 | FALSE |
| resilience:Agree->Disagree | Light touch users | 0.96 | 0.77 | 1.20 | FALSE |
| metro:Metro Area->Non-Metro Area | Light touch users | 0.96 | 0.78 | 1.19 | FALSE |
| can help others:Agree->Disagree | Light touch users | 0.97 | 0.78 | 1.20 | FALSE |
| race/ethnicity:white->other | Light touch users | 0.97 | 0.78 | 1.21 | FALSE |
| stigma:Disagree->Agree | Light touch users | 0.98 | 0.79 | 1.21 | FALSE |
| family emotional support:Agree->Disagree | Light touch users | 0.98 | 0.79 | 1.22 | FALSE |

| | | | | | |
|---|---|---|---|---|---|
| stress:Disagree->Agree | Light touch users | 0.98 | 0.79 | 1.22 | FALSE |
| belong to group:Agree->Disagree | Light touch users | 0.98 | 0.79 | 1.22 | FALSE |
| friend emotional support:Agree->Disagree | Light touch users | 1.00 | 0.80 | 1.23 | FALSE |
| worries social:0->1 | Light touch users | 1.00 | 0.80 | 1.24 | FALSE |
| neighborhood safety:Agree->Disagree | Light touch users | 1.00 | 0.80 | 1.24 | FALSE |
| any discrimination:0->1 | Light touch users | 1.00 | 0.80 | 1.24 | FALSE |
| confidence to achieve:Agree->Disagree | Light touch users | 1.00 | 0.81 | 1.25 | FALSE |
| live with caregivers:No->Yes | Light touch users | 1.01 | 0.81 | 1.26 | FALSE |
| physical health:Good->Poor | Light touch users | 1.02 | 0.82 | 1.27 | FALSE |
| gender:Woman->Another | Light touch users | 1.02 | 0.82 | 1.27 | FALSE |
| hard express:Disagree->Agree | Light touch users | 1.02 | 0.82 | 1.27 | FALSE |
| feel accomplishment:Agree->Disagree | Light touch users | 1.03 | 0.82 | 1.28 | FALSE |
| belong peers:Agree->Disagree | Light touch users | 1.03 | 0.83 | 1.28 | FALSE |
| symptoms depression/anxiety:No->Yes | Light touch users | 1.03 | 0.83 | 1.28 | FALSE |
| race/ethnicity:white->asian | Light touch users | 1.03 | 0.83 | 1.28 | FALSE |
| family accepts:Agree->Disagree | Light touch users | 1.03 | 0.83 | 1.28 | FALSE |

| | | | | | |
|---|---|---|---|---|---|
| neighbors look out each other:Agree->Disagree | Light touch users | 1.04 | 0.84 | 1.29 | FALSE |
| government assistance:Assistance->No Assistance | Light touch users | 1.05 | 0.85 | 1.30 | FALSE |
| mental health struggles:no_minimal_mh_struggles->mh_struggles_daily_impact | Light touch users | 1.06 | 0.86 | 1.32 | FALSE |
| gender:Woman->Man | Light touch users | 1.07 | 0.87 | 1.33 | FALSE |
| meaning in life:Agree->Disagree | Light touch users | 1.08 | 0.87 | 1.35 | FALSE |
| bullied:Disagree->Agree | Light touch users | 1.34 | 1.05 | 1.70 | TRUE |
| worries social:0->1 | Perpetually Plugged-In | 0.94 | 0.80 | 1.11 | FALSE |
| government assistance:Assistance->No Assistance | Perpetually Plugged-In | 0.95 | 0.80 | 1.12 | FALSE |
| live with caregivers:No->Yes | Perpetually Plugged-In | 0.97 | 0.83 | 1.15 | FALSE |
| belong peers:Agree->Disagree | Perpetually Plugged-In | 0.98 | 0.83 | 1.15 | FALSE |
| family accepts:Agree->Disagree | Perpetually Plugged-In | 0.98 | 0.83 | 1.16 | FALSE |
| worries global:0->1 | Perpetually Plugged-In | 0.99 | 0.84 | 1.17 | FALSE |
| symptoms depression/anxiety:No->Yes | Perpetually Plugged-In | 0.99 | 0.84 | 1.17 | FALSE |
| metro:Metro Area->Non-Metro Area | Perpetually Plugged-In | 0.99 | 0.84 | 1.17 | FALSE |
| physical health:Good->Poor | Perpetually Plugged-In | 1.00 | 0.84 | 1.17 | FALSE |
| belong to group:Agree->Disagree | Perpetually Plugged-In | 1.00 | 0.85 | 1.18 | FALSE |

| | | | | | |
|---|---|---|---|---|---|
| stress:Disagree->Agree | Perpetually Plugged-In | 1.00 | 0.85 | 1.18 | FALSE |
| can help others:Agree->Disagree | Perpetually Plugged-In | 1.01 | 0.85 | 1.19 | FALSE |
| gender:Woman->Man | Perpetually Plugged-In | 1.01 | 0.86 | 1.19 | FALSE |
| gender:Woman->Another | Perpetually Plugged-In | 1.01 | 0.86 | 1.19 | FALSE |
| neighborhood safety:Agree->Disagree | Perpetually Plugged-In | 1.01 | 0.86 | 1.19 | FALSE |
| stigma:Disagree->Agree | Perpetually Plugged-In | 1.02 | 0.86 | 1.20 | FALSE |
| hard express:Disagree->Agree | Perpetually Plugged-In | 1.02 | 0.87 | 1.21 | FALSE |
| race/ethnicity:white->hispanic | Perpetually Plugged-In | 1.03 | 0.87 | 1.21 | FALSE |
| family emotional support:Agree->Disagree | Perpetually Plugged-In | 1.03 | 0.87 | 1.21 | FALSE |
| friend emotional support:Agree->Disagree | Perpetually Plugged-In | 1.03 | 0.87 | 1.22 | FALSE |
| confidant:Agree->Disagree | Perpetually Plugged-In | 1.04 | 0.88 | 1.23 | FALSE |
| meaning in life:Agree->Disagree | Perpetually Plugged-In | 1.04 | 0.88 | 1.23 | FALSE |
| mental health struggles:no_minimal_mh_struggles->mh_struggles_daily_impact | Perpetually Plugged-In | 1.05 | 0.89 | 1.23 | FALSE |
| race/ethnicity:white->black | Perpetually Plugged-In | 1.05 | 0.89 | 1.24 | FALSE |
| neighbors look out each other:Agree->Disagree | Perpetually Plugged-In | 1.05 | 0.89 | 1.24 | FALSE |
| feel accomplishment:Agree->Disagree | Perpetually Plugged-In | 1.07 | 0.90 | 1.26 | FALSE |

| | | | | | |
|---|---|---|---|---|---|
| life satisfaction:Satisfied->Dissatisfied | Perpetually Plugged-In | 1.07 | 0.91 | 1.26 | FALSE |
| race/ethnicity:white->asian | Perpetually Plugged-In | 1.07 | 0.91 | 1.26 | FALSE |
| race/ethnicity:white->other | Perpetually Plugged-In | 1.07 | 0.91 | 1.27 | FALSE |
| confidence to achieve:Agree->Disagree | Perpetually Plugged-In | 1.11 | 0.94 | 1.31 | FALSE |
| age:15-17->18-24 | Perpetually Plugged-In | 1.16 | 0.98 | 1.37 | FALSE |
| LGBTQ+:Not LGBTQ->LGBTQ | Perpetually Plugged-In | 1.20 | 1.02 | 1.42 | TRUE |
| resilience:Agree->Disagree | Perpetually Plugged-In | 1.35 | 1.14 | 1.60 | TRUE |
| any discrimination:0->1 | Perpetually Plugged-In | 1.38 | 1.17 | 1.63 | TRUE |
| any ACE:0->1 | Perpetually Plugged-In | 1.43 | 1.21 | 1.68 | TRUE |
| emotional regulation:Disagree->Agree | Perpetually Plugged-In | 3.18 | 2.68 | 3.78 | TRUE |
| bullied:Disagree->Agree | Perpetually Plugged-In | 3.23 | 2.70 | 3.86 | TRUE |
| any ACE:0->1 | Positive engagers | 0.52 | 0.42 | 0.64 | TRUE |
| age:15-17->18-24 | Positive engagers | 0.84 | 0.69 | 1.03 | FALSE |
| LGBTQ+:Not LGBTQ->LGBTQ | Positive engagers | 0.89 | 0.72 | 1.09 | FALSE |
| emotional regulation:Disagree->Agree | Positive engagers | 0.91 | 0.74 | 1.12 | FALSE |
| stigma:Disagree->Agree | Positive engagers | 0.95 | 0.78 | 1.17 | FALSE |
| live with caregivers:No->Yes | Positive engagers | 0.95 | 0.78 | 1.17 | FALSE |
| worries global:0->1 | Positive engagers | 0.96 | 0.79 | 1.18 | FALSE |
| stress:Disagree->Agree | Positive engagers | 0.97 | 0.79 | 1.18 | FALSE |
| belong peers:Agree->Disagree | Positive engagers | 0.97 | 0.79 | 1.19 | FALSE |

| | | | | | |
|---|---|---|---|---|---|
| worries social:0->1 | Positive engagers | 0.97 | 0.79 | 1.19 | FALSE |
| physical health:Good->Poor | Positive engagers | 0.97 | 0.79 | 1.19 | FALSE |
| hard express:Disagree->Agree | Positive engagers | 0.97 | 0.79 | 1.19 | FALSE |
| meaning in life:Agree->Disagree | Positive engagers | 0.97 | 0.79 | 1.19 | FALSE |
| friend emotional support:Agree->Disagree | Positive engagers | 0.98 | 0.80 | 1.21 | FALSE |
| family accepts:Agree->Disagree | Positive engagers | 0.99 | 0.81 | 1.21 | FALSE |
| mental health struggles:no_minimal_mh_struggles->mh_struggles_daily_impact | Positive engagers | 0.99 | 0.81 | 1.21 | FALSE |
| metro:Metro Area->Non-Metro Area | Positive engagers | 0.99 | 0.81 | 1.22 | FALSE |
| resilience:Agree->Disagree | Positive engagers | 1.00 | 0.81 | 1.22 | FALSE |
| race/ethnicity:white->hispanic | Positive engagers | 1.00 | 0.81 | 1.22 | FALSE |
| belong to group:Agree->Disagree | Positive engagers | 1.00 | 0.81 | 1.22 | FALSE |
| can help others:Agree->Disagree | Positive engagers | 1.00 | 0.81 | 1.22 | FALSE |
| life satisfaction:Satisfied->Dissatisfied | Positive engagers | 1.00 | 0.81 | 1.23 | FALSE |
| feel accomplishment:Agree->Disagree | Positive engagers | 1.00 | 0.82 | 1.23 | FALSE |
| gender:Woman->Another | Positive engagers | 1.00 | 0.82 | 1.23 | FALSE |
| family emotional support:Agree->Disagree | Positive engagers | 1.01 | 0.82 | 1.24 | FALSE |
| neighbors look out each other:Agree->Disagree | Positive engagers | 1.01 | 0.83 | 1.24 | FALSE |
| race/ethnicity:white->other | Positive engagers | 1.02 | 0.83 | 1.25 | FALSE |
| confidant:Agree->Disagree | Positive engagers | 1.02 | 0.83 | 1.24 | FALSE |
| race/ethnicity:white->asian | Positive engagers | 1.02 | 0.84 | 1.25 | FALSE |
| symptoms depression/anxiety:No->Yes | Positive engagers | 1.02 | 0.84 | 1.26 | FALSE |
| confidence to achieve:Agree->Disagree | Positive engagers | 1.03 | 0.84 | 1.26 | FALSE |
| gender:Woman->Man | Positive engagers | 1.03 | 0.84 | 1.27 | FALSE |

| | | | | | |
|---|---|---|---|---|---|
| race/ethnicity:white->black | Positive engagers | 1.05 | 0.86 | 1.29 | FALSE |
| neighborhood safety:Agree->Disagree | Positive engagers | 1.07 | 0.87 | 1.31 | FALSE |
| government assistance:Assistance->No Assistance | Positive engagers | 1.14 | 0.93 | 1.39 | FALSE |
| any discrimination:0->1 | Positive engagers | 1.16 | 0.94 | 1.42 | FALSE |
| bullied:Disagree->Agree | Positive engagers | 2.42 | 1.95 | 2.99 | TRUE |

Supplementary 4: Counterfactual interventions on factors downstream to segment

*Relative to remaining Positive Engagers*

| outcome | segment | intervention | Downstream impact | OR | Lower CI | Upper CI | sig |
|---|---|---|---|---|---|---|---|
| belong | Light touch users | social media segment:Positive engagers->Light touch users | Agree->Disagree | 1.23 | 1.01 | 1.50 | TRUE |
| belong | Practical navigators | social media segment:Positive engagers->Practical navigators | Agree->Disagree | 1.45 | 1.20 | 1.76 | TRUE |
| belong | Perpetually Plugged-In | social media segment:Positive engagers->Perpetually Plugged-In | Agree->Disagree | 1.70 | 1.41 | 2.04 | TRUE |
| belong | Burned-Out Browsers | social media segment:Positive engagers->Burned-Out Browsers | Agree->Disagree | 2.56 | 2.14 | 3.07 | TRUE |
| symptoms depression/anxiety | Practical navigators | social media segment:Positive engagers->Practical navigators | No->Yes | 0.68 | 0.56 | 0.82 | TRUE |
| symptoms depression/anxiety | Light touch users | social media segment:Positive engagers->Light touch users | No->Yes | 0.93 | 0.78 | 1.11 | FALSE |
| symptoms depression/anxiety | Burned-Out Browsers | social media segment:Positive engagers->Burned-Out Browsers | No->Yes | 1.46 | 1.24 | 1.73 | TRUE |
| symptoms depression/anxiety | Perpetually Plugged-In | social media segment:Positive engagers->Perpetually Plugged-In | No->Yes | 2.25 | 1.92 | 2.64 | TRUE |

| | | | | | | | |
|---|---|---|---|---|---|---|---|
| can help others | Light touch users | social media segment:Positive engagers->Light touch users | Agree->Disagree | 1.20 | 0.95 | 1.52 | FALSE |
| can help others | Practical navigators | social media segment:Positive engagers->Practical navigators | Agree->Disagree | 1.45 | 1.15 | 1.81 | TRUE |
| can help others | Perpetually Plugged-In | social media segment:Positive engagers->Perpetually Plugged-In | Agree->Disagree | 1.50 | 1.21 | 1.86 | TRUE |
| can help others | Burned-Out Browsers | social media segment:Positive engagers->Burned-Out Browsers | Agree->Disagree | 1.55 | 1.25 | 1.92 | TRUE |
| mental health struggles | Light touch users | social media segment:Positive engagers->Light touch users | no_minimal_mh_struggles->mh_struggles_daily_impact | 0.71 | 0.61 | 0.83 | TRUE |
| mental health struggles | Practical navigators | social media segment:Positive engagers->Practical navigators | no_minimal_mh_struggles->mh_struggles_daily_impact | 0.96 | 0.83 | 1.11 | FALSE |
| mental health struggles | Burned-Out Browsers | social media segment:Positive engagers->Burned-Out Browsers | no_minimal_mh_struggles->mh_struggles_daily_impact | 1.94 | 1.70 | 2.23 | TRUE |
| mental health struggles | Perpetually Plugged-In | social media segment:Positive engagers->Perpetually Plugged-In | no_minimal_mh_struggles->mh_struggles_daily_impact | 2.79 | 2.44 | 3.19 | TRUE |
| stigma | Practical navigators | social media segment:Positive engagers->Practical navigators | Disagree->Agree | 0.95 | 0.84 | 1.09 | FALSE |
| stigma | Light touch users | social media segment:Positive engagers->Light touch users | Disagree->Agree | 1.01 | 0.89 | 1.15 | FALSE |

| | | | | | | | |
|---|---|---|---|---|---|---|---|
| stigma | Burned-Out Browsers | social media segment:Positive engagers->Burned-Out Browsers | Disagree->Agree | 2.11 | 1.87 | 2.39 | TRUE |
| stigma | Perpetually Plugged-In | social media segment:Positive engagers->Perpetually Plugged-In | Disagree->Agree | 2.54 | 2.25 | 2.87 | TRUE |
| stress | Practical navigators | social media segment:Positive engagers->Practical navigators | Disagree->Agree | 0.60 | 0.52 | 0.70 | TRUE |
| stress | Light touch users | social media segment:Positive engagers->Light touch users | Disagree->Agree | 0.79 | 0.69 | 0.91 | TRUE |
| stress | Burned-Out Browsers | social media segment:Positive engagers->Burned-Out Browsers | Disagree->Agree | 1.20 | 1.06 | 1.37 | TRUE |
| stress | Perpetually Plugged-In | social media segment:Positive engagers->Perpetually Plugged-In | Disagree->Agree | 2.13 | 1.88 | 2.41 | TRUE |

### Supplementary 5 Survey Measures

#### Mental health and wellbeing

Depressive and anxiety symptoms were assessed using the PHQ-2 and GAD-2 screening items (Kroenke et al., 2003; Kroenke et al., 2007). Perceived mental health burden was additionally assessed using a study-specific item indicating whether respondents had struggled with their mental health in the past year. Life satisfaction was measured using a single item from the National Health Interview Survey. Additional indicators of wellbeing, including meaning in life and sense of accomplishment, were assessed using items adapted from established flourishing frameworks (Huppert & So, 2013). A global measure of perceived stress was captured using an item from the Perceived Stress Scale (Cohen et al., 1983).

#### Social support and belonging

Perceived social support was assessed using items adapted from the Multidimensional Scale of Perceived Social Support (Canty-Mitchell & Zimet, 2000), capturing support from family and friends as well as the presence of a confidant. Family acceptance and peer belonging were measured using items aligned with the Milwaukee Youth Belongingness Scale (Slaten et al., 2019). Indicators of social contribution and integration were adapted from measures used in the Child Development Supplement (CDS–II) of the Panel Study of Income Dynamics (PSID) (Keyes, 2006).

#### Emotional regulation and resilience

Emotional regulation and resilience were assessed using selected items adapted from the Adolescent Resilience Questionnaire (Anderson et al., 2020), including difficulty managing negative emotions, emotional expression, and confidence in achieving goals. Ability to recover from setbacks was adapted from established flourishing frameworks (Huppert & So, 2013).

#### Adversity and social stressors

Exposure to adverse childhood experiences (ACEs) was assessed using a set of items adapted from the National Health Interview Survey stressful life events module, capturing experiences of unmet basic needs, household dysfunction (including parental substance use, mental illness, incarceration, and emotional abuse), and exposure to violence in the home or neighborhood. A composite ACE indicator was constructed to reflect whether respondents had experienced any of these adversities. Exposure to discrimination was assessed using items capturing experiences of unfair treatment based on sexual orientation, gender identity, race/ethnicity (including skin color, language, or culture), country of origin, and religion. Items were adapted and expanded from measures derived from the Perceptions of Racism in Children and Youth instrument (Pachter et al., 2010). A binary indicator was constructed to reflect whether respondents reported experiencing any form of identity-based discrimination.  Experiences of bullying and mental health stigma were assessed using study-specific items.

#### Environmental, structural and individual context

Perceived neighborhood safety and social cohesion were assessed using study-specific items adapted from existing measures (Anderson et al., 2021). Additional variables captured physical health, financial assistance, and demographic characteristics.

Social media experiences and behaviors

Social media experiences were assessed by a set of study-specific items, informed by prior literature and cognitively tested, capturing both negative experiences (e.g., social comparison, appearance concerns) and positive uses (e.g., identity exploration, connection, emotional support). Frequency and intensity of digital device use and attempts to reduce use were also assessed.
